# ACT 2 Clinical Trial Design: Acceptability and Feasibility of Combination Treatment for Cervical Precancer Among South African Women Living with HIV

**DOI:** 10.1101/2025.08.12.25333540

**Authors:** Carla J. Chibwesha, Katie R. Mollan, Nicholas S. Teodoro, Jessica R. Keys, Chuning Liu, Masangu Mulongo, Sibusisiwe Gumede, Tafadzwa Pasipamire, Mark Faesen, Lisa Rahangdale

## Abstract

**Background:** Global efforts to eliminate cervical cancer currently focus on expanding access to HPV vaccination and cervical screening. However, these efforts lack equal investment in treatment of precancerous cervical intraepithelial neoplasia grade 2/3 (CIN2/3), particularly among women with HIV (WWH). This gap leaves a generation of WWH at high risk of persistent/recurrent CIN2/3 and progression to cervical cancer, as standard surgical treatments alone are far less effective in this population.

**Methods and Design:** After randomization, WWH with CIN2/3 (N=180) are randomly allocated to undergo Loop Electrosurgical Excision Procedure (LEEP) followed by self-administration of 8 doses of intravaginal 5% 5-fluorouracil (5FU) or placebo cream (one dose every other week for 16 weeks). Women will be followed for 24 weeks. The primary outcomes are acceptability and feasibility (safety, tolerability, adherence, retention). Secondary outcomes include (a) regression of cervical disease to CIN1 or normal histology and (b) clearance of the hrHPV genotype(s) identified in baseline cervical ThinPrep samples.

**Discussion:** We hypothesize that topical 5FU, a widely available low-cost generic drug, can be repurposed as an adjuvant treatment for CIN2/3 to be self-administered after surgical excision (or ablation) to reduce the risk of persistent/recurrent CIN2/3 and progression to cervical cancer among WWH. In preparation for a large-scale trial to test the effectiveness of combination treatment for CIN2/3 in WWH, we are conducting a Phase 2b feasibility trial of LEEP combined with intravaginal 5FU.

**Trial Registration:** ClinicalTrials.gov, NCT05413811.

## INTRODUCTION

Cervical cancer remains the second most common cancer among women worldwide, and over 85% of the global burden of this disease occurs in the developing world. Women with HIV (WWH) are at increased risk of HPV infection, with rates as high as 45% - 90%.^1–3^ Despite being preventable, cytologic abnormalities, cervical precancer (high-grade cervical intraepithelial neoplasia [CIN2/3]), and invasive cervical cancer also occur more frequently in WWH. ^2,4^

Current management of CIN2/3 is based on surgical therapy alone, which is less effective in WWH.^5^ Surgical therapy for cervical precancer can be excisional (e.g., cold knife conization, loop electrosurgical excision procedure [LEEP]) or ablative (e.g., cryotherapy, thermocoagulation, laser ablation).^6^ Excisional treatment requires more surgical skill, but is more appropriate for larger lesions and has the advantage of producing a biopsy specimen for histologic review.^7^ Excision is also the standard of care for treatment of CIN2/3 in South Africa.^8^

Although surgical therapy is highly successful in women without HIV,^9,10^ cure rates for CIN2/3 are significantly lower in HIV-infected women.^11,12^ Three recent African studies, enrolling a combined 791 HIV-infected women on antiretroviral therapy (ART), confirm a high risk (19-37%) of recurrent CIN2/3 among women treated with either LEEP or cryotherapy at 12-24 months.^13,14^ These findings are consistent with a prior U.S. study in which treatment failure occurred in 55% of women with CIN2/3 (n=75) over a 6-year period.^11^ By comparison, LEEP treatment failure occurs in only 5-10% of women who are not HIV-infected.^9^

The increase in cervical disease persistence or recurrence among WWH is thought to derive from residual CIN2/3 after excision,^15^ re-infection with another high-risk HPV(hrHPV) type, and/or reactivation of underlying hrHPV infection in the genital tract.^16^ WWH could therefore benefit from effective adjuvant therapy following surgery for CIN2/3.

Multiple observational studies support the efficacy of topical 5FU for treatment of HPV-related diseases (e.g., genital warts, vulvar and vaginal precancer).^17–19^ Intravaginal 5FU was initially studied in the in U.S. as adjuvant therapy for cervical disease in HIV-infected women following surgical treatment for CIN2/3.^20^ Women self-applied 2 grams of 5% intravaginal 5FU every 2 weeks for up to 6 months. Histologic outcomes were assessed over 18 months. Participants were relatively immune-compromised; 35% had CD4 counts of <200 cells/mm^3^ and half were ART-naïve. Compared to women in the observation arm, women treated with 5FU were less likely to experience CIN2/3 recurrence.^20^

Rahangdale et al. hypothesized that a similar low-frequency dosing schedule of intravaginal 5FU (2g every 2 weeks) could serve as primary treatment for CIN2 women without HIV.^21^ Their trial enrolled immunocompetent women and therapy was shortened to 16 weeks, consistent with other HPV-related medical management studies.^22–25^ Sixty healthy women were randomly assigned to self-apply 5FU every 2 weeks for 16 weeks (8 total doses) versus observation. Compared to observation, women in the 5FU treatment arm were more likely to experience disease regression to CIN1 or normal histology (93% 5FU versus 56% control).^21^ Additionally, the composite outcome of histology, Pap smear, and hrHPV result was more likely to be normal in the 5FU treatment group (relative risk: 2.25; 95% CI: 1.05 to 5.09). With respect to adverse events (AEs), 48% of women reported grade 1 events (e.g., discharge, burning/irritation, spotting). None of the participants experienced symptoms that caused interference with their usual activities and 83% reported feeling satisfied with the use of 5FU cream.^21^

## METHODS AND DESIGN

### Aim and Objectives

The overall aim of our research is to reduce the risk of persistent/recurrent CIN2/3 and progression to cervical cancer among WWH by studying combination treatment approaches for CIN2/3 in this high-risk population. This project was conducted in two phases: a formative study followed by a clinical trial. The primary objective of our clinical trial is to determine the acceptability and feasibility of combination treatment of CIN2/3 (surgical excision followed by adjuvant 5FU) among WWH. Our secondary objectives are related to the efficacy of combination treatment for CIN2/3. At week 24, we will assess (a) regression of cervical disease to CIN1 or normal histology and (b) clearance of the hrHPV genotype(s) identified in baseline cervical ThinPrep samples.

### Study Design, Randomization, and Masking

To test our hypothesis that combination treatment for CIN2/3 (i.e., surgical excision followed by adjuvant 5FU) will be safe and well tolerated, we are conducting a Phase 2b feasibility trial. The trial, known as “ACT 2” is a double-blind, randomized, and placebo controlled. Briefly, WWH with CIN2/3 confirmed on LEEP will be randomly assigned (1:1) to receive 8 doses of intravaginal 5FU cream or placebo cream (once every 2 weeks). Participants are followed for 24 weeks to assess acceptability and feasibility (safety, tolerability, adherence, and retention) of the combined treatment approach (Figure 1).

**Figure 1.**
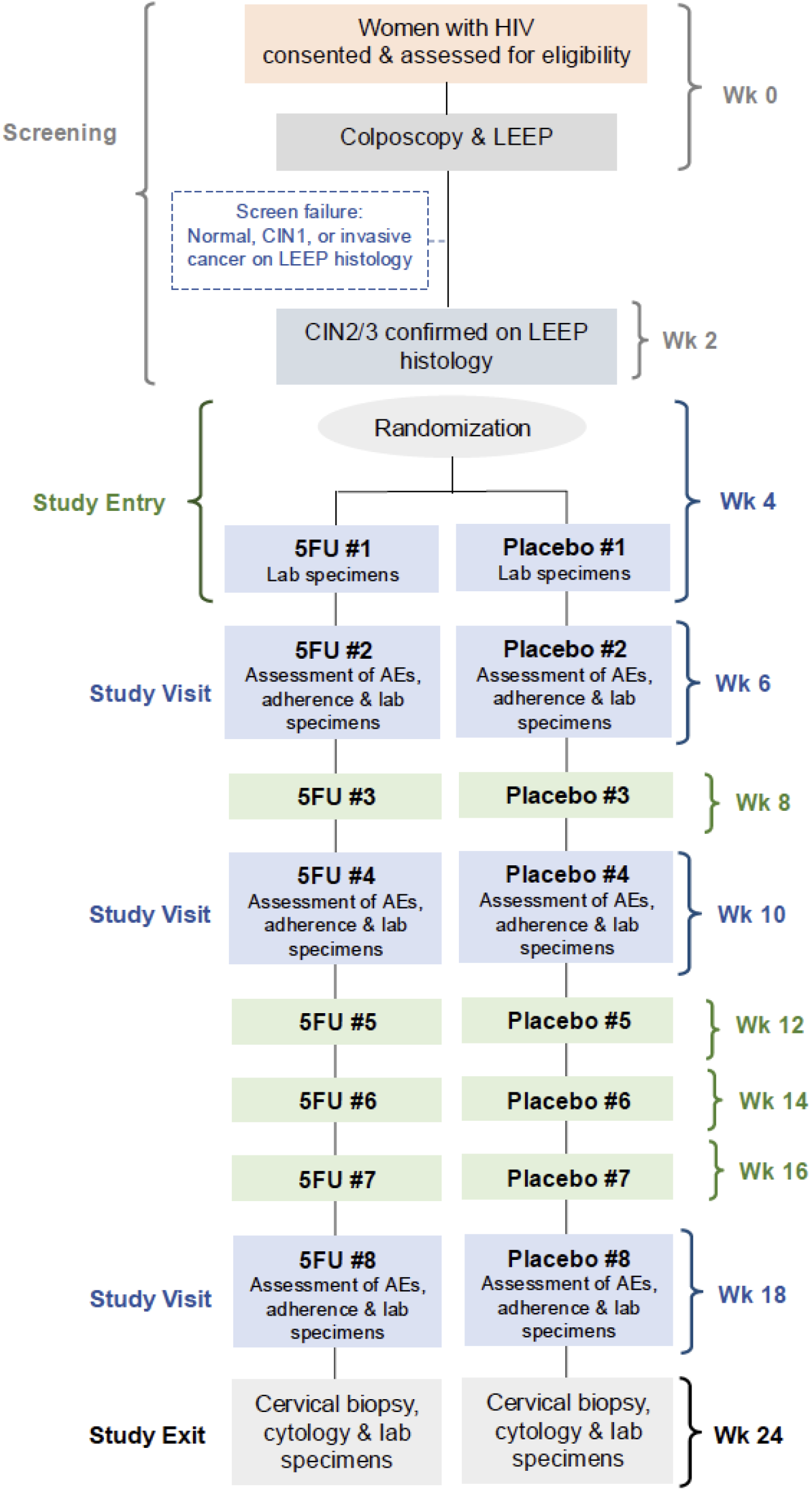
Trial Schema.

The randomization schedule was generated by the study statistician in SAS version 9.4 using a random number generator and a blocked randomization design. The block size(s) will be kept concealed from the study team throughout the trial. Randomization IDs were assigned in a concealed manner using a central, web-based REDCap utility. An unmasked study pharmacist used the randomization schedule to provide the assigned 5FU or placebo treatment.

### Ethical Approval and Clinical Trial Registration

This protocol has been reviewed and approved by the University of Witwatersrand Human Research Ethics Committee (201122) and the University of North Carolina Institutional Review Board (20-3565). The trial is publicly registered on the South African National Clinical Trials Register and with U.S. National Library of Medicine at ClinicalTrials.gov (NCT05413811).

### Study Setting

ACT 2 is being conducted in Johannesburg, South Africa. WWH are recruited from the cervical cancer prevention program at an HIV clinic located in a large public-sector teaching hospital. The research team includes national leaders in cervical cancer prevention efforts and seasoned clinical trialists.^13,26-39^

South Africa is home to more than 5 million WWH and has the world’s largest public-sector HIV treatment programs.^40^ Despite substantial local and international investments in HIV, cervical cancer remains a leading cause of cancer deaths in the country^6,10^ due to suboptimal screening coverage rates and fragmented linkages to colposcopy and LEEP treatment services.^41–44^

### Study Population

The trial opened in March 2023 under protocol version 3, which included the following inclusion criteria: women with (a) confirmed HIV-1 infection; (b) age ≥ 18 years; (c) on ART for at least 90 days prior to enrollment; and (d) biopsy-proven CIN2/3 within the preceding 120 days. To be eligible, women should also be willing and able to provide written informed consent.

An amendment to the inclusion criteria was made in April 2023 and under protocol v4; women who met the following eligibility criteria are included: (a) confirmed HIV-1 infection; (b) age ≥ 18 years; (c) on ART for at least 60 days prior to enrollment; and (d) eligible for LEEP (i.e., those with a cervical biopsy demonstrating CIN2/3 or HSIL cytology on Pap smear within the preceding 12 months).

Under both protocol v3 and v4, women are excluded from participation if they are (a) pregnant, breastfeeding, or intend to become pregnant within 180 days of enrollment; (b) have an active sexually transmitted infection (STI; women may participate once treated); (c) have a surgically absent cervix; (d) have a history of anogenital (cervical, vaginal, vulvar, or anal) cancer or a biopsy suspicious for cervical cancer; or (e) have a medical comorbidity that would interfere with study participation.

Because intravenous 5FU has been shown to be a teratogen in animal models and is classified as a Pregnancy Category X drug, study participants will be required to use dual contraception (condoms plus hormonal contraception, an intrauterine device, implant, or sterilization) during the study.^45^ Hormonal contraception and condoms are provided free of charge to all study participants.

### Study Intervention

LEEP is provided in accordance with the standard of care. 5FU cream (trade name Efudix, generic supplied from Mylan, South Africa) is supplied locally and held in our research pharmacy. The study placebo cream is an inert emollient that mimics the consistency and coloring of the 5FU cream. Vaginal applicators pre-filled with 2g of 5% 5FU cream are distributed to participants to self-apply the cream intravaginally once every 2 weeks for a total of 8 doses. The placebo cream is also supplied to participants in pre-filled applicators and has identical dosing instructions.

Educational materials for the trial were co-developed with participants during our formative study.^46^ These materials include step-by-step instructions on how to use the 5FU/placebo cream (Supplemental File 1).

### Study Procedures

WWH who are clinically eligible for LEEP (i.e., those with a cervical biopsy demonstrating CIN2/3 or HSIL cytology on Pap smear within the preceding 12 months) are screened for participation. Those wishing to participate provide written informed consent and are assessed for eligibility, which will include medical history and concurrent medication use, urine pregnancy testing, pelvic examination, STI testing for *C. trachomatis, N. gonorrhoeae, T. vaginalis* and screening for syphilis.

At their screening visit (week 0), women undergo colposcopy and LEEP. The excised cervical tissue is submitted for histopathology review to confirm eligibility enrollment in the trial. Additional baseline specimens collected at the screening visit include a ThinPrep sample for HPV testing and blood samples for CD4 and HIV-1 plasma viral load testing. A cervicovaginal lavage (CVL) sample and vaginal swab are also collected for storage and future testing (Table 1).

**Table 1.**
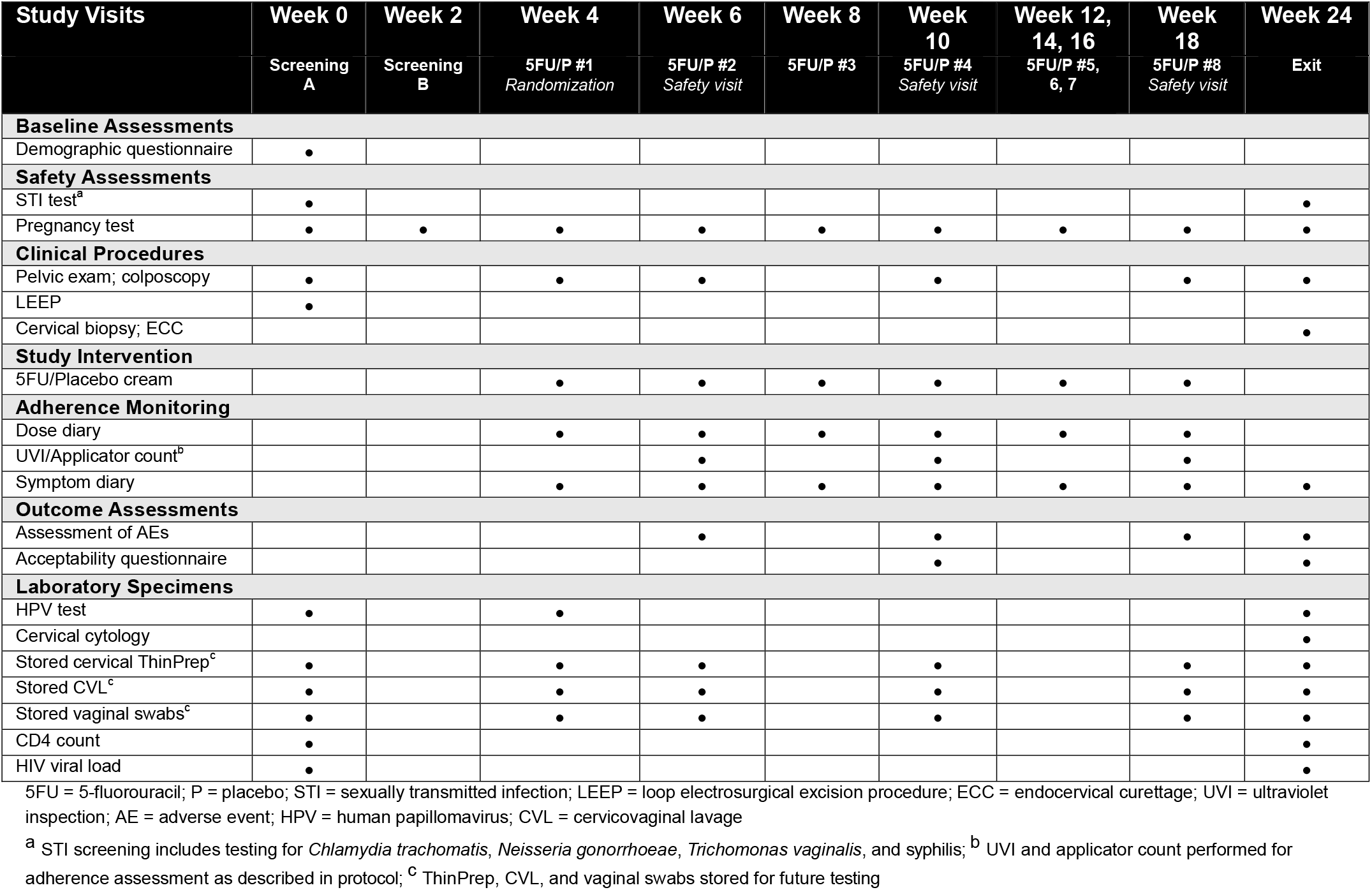
Schedule of Events.

Women return at week 2 to receive their LEEP histology results and confirm eligibility for participation. At week 4 (enrollment visit), those with CIN2/3 confirmed on LEEP histology are randomly assigned (1:1) to receive 8 doses of intravaginal 5FU or placebo once every 2 weeks. The 5FU/placebo treatment begins at week 4 to allow time for women to heal from their LEEP.

Additionally, at week 4, women undergo pregnancy testing, pelvic examination, and colposcopy, and receive step-by-step instructions for 5FU/placebo cream use and a demonstration using a pelvic model. Women apply the first dose of cream in the study clinic and observed for immediate adverse events. They are also supplied with pre-filled vaginal applicators of 2g doses of 5FU/placebo cream and asked to apply 2g of the cream at home every 2 weeks (8 doses total, with additional at-home doses at weeks 6, 8, 10, 12, 14, 16, and 18). Women are also provided with symptom and dose diaries, condoms, and urine pregnancy tests to be used prior to applying the 5FU/placebo at home. At week 4, we also collect a cervical ThinPrep sample for HPV testing, as well as CVL and a vaginal swab for storage.

Regardless of study arm, women return for additional study visits at weeks 6, 10, 18, and 24. Study procedures during the visits at week 6, 10, and 18 include pregnancy testing, pelvic examination, and colposcopy to assess possible AEs. At each of these visits, we collect cervical ThinPrep, CVL, and a vaginal swab for sample for storage. Adherence is also reinforced at weeks 6, 10, and 18.

At week 24 (exit visit), women undergo colposcopy, cervical biopsy, and, if clinically indicated, endocervical curettage (ECC). STI testing is performed at this visit and we also collect liquid-based cytology, cervical ThinPrep for HPV testing, and blood samples for CD4 and HIV-1 plasma viral load testing. As at prior visits, CVL and vaginal swab are collected for storage and future testing.

### Gynecologic Procedures

Our research team performs all gynecologic procedures in their study clinic and is blinded to study arm assignment. At each study visit, women undergo pelvic examination of the vulva, vagina, cervix, and perineum as part of the clinical assessment for STIs and AEs. During colposcopy 5% acetic acid is be applied to the cervix and digital images of the cervical transformation zone are obtained. These images will be externally reviewed as a quality assurance measure to minimize inter-observer variability in clinical assessment and biopsy location.

All participants undergo LEEP during their screening visit. After paracervical nerve block with injected lidocaine, a wire loop is used to excise abnormal-appearing tissue in the cervical transformation zone in accordance with the standard of care. Tissue samples are sent to our research laboratory for histologic evaluation and, if clinically indicated, immunohistochemical (IHC) staining. Similarly, at week 24, all participants undergo colposcopically-directed, 4-quadrant biopsy and ECC to adequately evaluate the cervical transformation zone.^47,48^ Tissue samples will be sent for histologic evaluation and IHC staining, if clinically indicated.

Liquid-based cytology (LBC) specimens (Pap smears) and cervical specimens for HPV testing/storage are both collected using a Cervex-Brush (Rovers Medical Devices, Oss, Netherlands), while vaginal swab samples are collected using polyester-tipped swab used to swab the walls of vagina. CVL is performed by aiming a continuous stream of 10mL of phosphate-buffered saline directly at and into the cervical opening to bathe the endocervix and ectocervix. The fluid pool is then aspirated from the posterior fornix.

### Laboratory Testing

Central review of both histology and cytology specimens is performed by pathologists blinded to the study arm assignment. Histology slides are prepared and stained by a histotechnologist, read by a histopathologist, and reported in accordance 2012 Lower Anogenital Squamous Terminology (LAST) guidelines.^49,50^ Similarly, initial screening of the cytology specimens is performed by a cytotechnologist, with all smears to be reviewed by a cytopathologist. For both histology and cytology, consultation and consensus review is used to resolve discrepancies

HPV testing of cervical ThinPrep samples is performed using the GeneXpert system (GeneXpert; Cepheid, Sunnyvale, CA), which employs a cartridge-based real-time PCR system and reports five separate results: (a) HPV16, (b) HPV18/45, (c) HPV 31/33/35/52/58, (d) HPV51/59, (e) HPV39/68/56/66. Sensitivity and specificity of Xpert HPV in both general screening populations and WWH is comparable to other commercially available HPV tests.^51,52^

STI testing for *N. gonnorhea, C. trachomatis*, and *T. vaginalis* using vaginal samples is also performed using the GeneXpert system (GeneXpert; Cepheid, Sunnyvale, CA).^53–56^ Rapid testing for syphilis will be performed using Syphilis BD Macro-Vue (Abbott, Abbott Park IL), a screening assay for *T. pallidum* antibodies.^57^ Women diagnosed with STIs receive treatment in accordance with the local standard of care. Their partners also receive empiric STI treatment.

To assess participants’ immune status, CD4+T-cell counts are determined by flow cytometry performed on whole blood samples using the FACSCanto Clinical Flow Cytometry System (BD Biosciences, Franklin Lakes, New Jersey). Plasma HIV viral load is measured using the Abbott RealTime HIV-1 Assay (Abbott m2000 system, Abbott Laboratories, Abbott Park, Illinois). The range of detection for the HIV assay is 40 to 10,000,000 copies/mL. All testing is performed in accordance with the manufacturer’s instructions.

Residual plasma and cervical ThinPrep samples are aliquoted and stored at −70°C (or below) for future HPV, DNA methylation, microbiome, and related biomarker analyses. CVL samples are aliquoted and stored at −70°C (or below) for future HIV viral load and cytokine testing. Vaginal samples are also aliquoted and stored at −70°C (or below) for future HPV and microbiome analyses. Lastly, FFPE tissue specimens are stored at ambient temperature for future HPV, DNA methylation, and related biomarker analyses.

### Assessment of Safety and Tolerability of the Study Intervention

Our trial adheres to National Institutes of Health (NIH) AE reporting guidelines. We identify AEs using NCI’s Common Terminology Criteria for Adverse Events (CTCAE) version 5.0 and the DAIDS Female Genital Grading Table for Use in Microbicide Studies v1.0. Participants use a symptom diary to self-report data on cream use and side effects. During study visits at weeks 4, 6, 10, 18, and 24, participants also undergo pelvic examination and colposcopy. All AEs noted during these visits are graded using the CTCAE and DAIDS grading tables.

We collect data on toxicities as outlined above and will report dose-limiting toxicities (DLT), defined as (a) any Grade 2 or greater AE that that is possibly, probably, or definitely related to study drug and results in a reduction to the number of doses or discontinuation the study drug, or (b) any Grade 1 AE of any genital lesion (e.g., blisters, ulcerations, or pustules) that is possibly, probably, or definitely related to study drug and results in results in a reduction to the number of doses or discontinuation the study drug.

### Assessment of Adherence to the Study Intervention

We will study adherence to intravaginal cream using a combination of data collection methods to limit the potential for recall and social desirability bias often introduced when data collection occurs at study visits rather than in real time.^58–60^ Thus, we measure adherence using 3 previously-validated methods: applicator counts,^61^ ultraviolet inspection (UVI),^62^ and self-report.^63^

Applicators containing 5FU/placebo cream will be placed into a resealable bag. Women are asked to return all used and unused applicators at follow-up visits, and these applicators are counted and recorded.^61^ Applicators are tested for evidence of vaginal insertion using a UV light. The sensitivity and specificity of the UVI approach (previously validated in South Africa) is 83% and 92%, respectively.^62^ Finally, self-reported data on cream use are collected using a dose diary.

### Outcome Measures

Our primary outcomes are acceptability and feasibility, incorporating assessments of safety, tolerability, adherence, and retention. Acceptability is measured at weeks 10 and 24 using a questionnaire with 7 Likert scale items. Feasibility (safety, tolerability, adherence, and retention) are assessed between weeks 4 and 24.

Our safety endpoint is the number and percentage of women in each study arm experiencing a grade 2 or higher AE, or any grade 1 genital AE (e.g., blisters, ulcerations, or pustules), that is possibly, probably, or definitely related to the study intervention. Our tolerability endpoint is number and percentage of women in each study arm unable or unwilling to apply at least 4 of 8 doses (50%) of the 5FU/placebo cream due to a DLT. For our adherence endpoint, three sources of adherence data (applicator counts, UVI, and self-report) are collected and the corresponding adherence rates will be reported. Intra-class correlation (ICC) is used to estimate within-woman agreement between the three data sources.^64^ UVI is considered the primary adherence measure. Finally, our retention endpoint is the number and percentage of women in each study arm who are retained in study follow-up over the 24-week study duration.

Our secondary outcomes are related to the efficacy of combination treatment for CIN2/3. At week 24, we will assess (a) regression of cervical disease to CIN1 or normal histology and (b) clearance of the hrHPV genotype(s) identified at baseline (week 0).

### Statistical Analysis Plan

Descriptive statistics will be used to (a) delineate the overall study sample that was recruited from a population of WWH in South Africa who are on ART and have a clinical indication for LEEP, and (b) assess for clinically meaningful imbalances between the randomly assigned study arms (see Supplemental File 2 for the Statistical Analysis Plan). For the primary acceptability and feasibility endpoints, our main analyses will include all participants who are study eligible, enrolled, randomized, and who receive at least one dose of 5FU or placebo (i.e., a modified intention-to-treat (ITT) approach). Additionally, per-protocol and ITT analyses will be conducted as detailed in Supplemental File 2.

#### Acceptability

We will describe the responses to each Likert scale item individually. To compare 5FU vs. placebo, an acceptability summary score will be calculated as a percentage (0 to 100%) of 0 to 28 possible Likert scale points, and the distribution of scores will be compared between randomization arms at weeks 10 and 24, separately, using a Wilcoxon rank-sum test and descriptive statistics. We will then estimate the proportion of women who reported 80% or higher acceptability summary score in each randomization arm, and a corresponding binomial 95% CI will be constructed for each study arm. Lastly, we will estimate the difference in proportions (5FU vs. Placebo) with a corresponding 95% CI.

#### Safety

We will report the number and percentage of women in each randomization arm experiencing a safety event. A Kaplan Meier (KM) estimate of safety event probability and a corresponding 95% CI will be constructed for each randomization arm at study week 24, and safety will be compared between the randomization arms using an estimated risk difference (5FU - placebo) with a corresponding 95% CI. The time origin is randomization (week 4).

#### Tolerability

We will report the number and percentage of women in each randomization arm unable or unwilling to apply at least 4 of 8 doses of the 5FU/placebo cream due to a DLT. A KM estimate of tolerability event probability and a corresponding 95% CI will be constructed for each randomization arm at study week 18, and tolerability will be compared between the randomization arms using a risk difference (5FU - placebo) with a corresponding 95% CI.

#### Adherence

For each of the three adherence measures, we will derive a binary endpoint classifying each woman as adherent if she administers at least 6 of 8 doses of 5FU/placebo. Within each randomization arm, we will report the number and percentage of women achieving this adherence threshold with a corresponding binomial 95% CI. The percentage of women who achieve adherent dosing will be compared between the randomization arms using an estimated difference in proportions with a corresponding 95% CI.

#### Retention

We will report the number and percentage of women in each randomization arm who are retained in study follow-up at each visit over the 24-week study duration. Week 24 retention will be estimated within each randomization arm with a corresponding binomial 95% CI. Week 24 retention will be compared between the 5FU and placebo arms using an estimated difference in proportions and corresponding 95% CI.^65^

#### Regression of cervical disease

All eligible participants will have CIN2/3 (confirmed by LEEP histology) measured at study week 0 (baseline for this analysis). We will report the number and percentage of women in each randomization arm who regress (improve) to CIN1 or normal at study week 24 with a corresponding binomial 95% CI. Regression of cervical disease will be compared between the randomization arms using an estimated difference in proportions and corresponding 95% CI.

#### Clearance of hrHPV

We will report the number and percentage of women in each randomization arm who demonstrate genotype-specific hrHPV clearance, defined as having none of the hrHPV types at week 24 that were originally present at baseline (week 0). A corresponding binomial 95% CI will be constructed. This composite measure of hrHPV type-specific clearance will be compared between the randomization arms using an estimated difference in proportions and corresponding 95% CI.

### Sample Size

We are enrolling 180 participants for 1:1 randomization to 5FU or placebo cream. *A priori*, we anticipate grade 2 or higher safety AEs and tolerability events to be uncommon,^21^ thus our sample size calculations first addressed the acceptability and adherence endpoints; then we calculated anticipated precision for the safety and tolerability endpoints. Precision and power calculations for secondary outcomes are included in the Supplemental File 2. We assume ≥90% of women will be retained in study follow-up, thus throughout our precision and power calculations assume 10% attrition over the course of the study.

To estimate the proportions of women reporting an 80% or higher acceptability score, overall and in each arm, we calculated a binomial 95% CI. We assume 80% of women will achieve this acceptability threshold at weeks 10 and 24, and thus enrolling 180 women will provide an anticipated margin of error of ±8.7% within arm (Table 2). Likewise, to estimate the proportions achieving adherence overall and within each arm, we calculated a binomial 95% CI. We assumed that 80% of women will achieve adequate adherence (i.e., 6 of 8 doses). To measure this proportion with a margin of error <10% (anticipated ±8.7%) within arm, we will enroll 180 women. If we observe 90% retention over 24 weeks the 95% CI precision for retention will be (0.85, 0.94) pooled over arms and (0.82, 0.95) within arm.

**Table 2.**
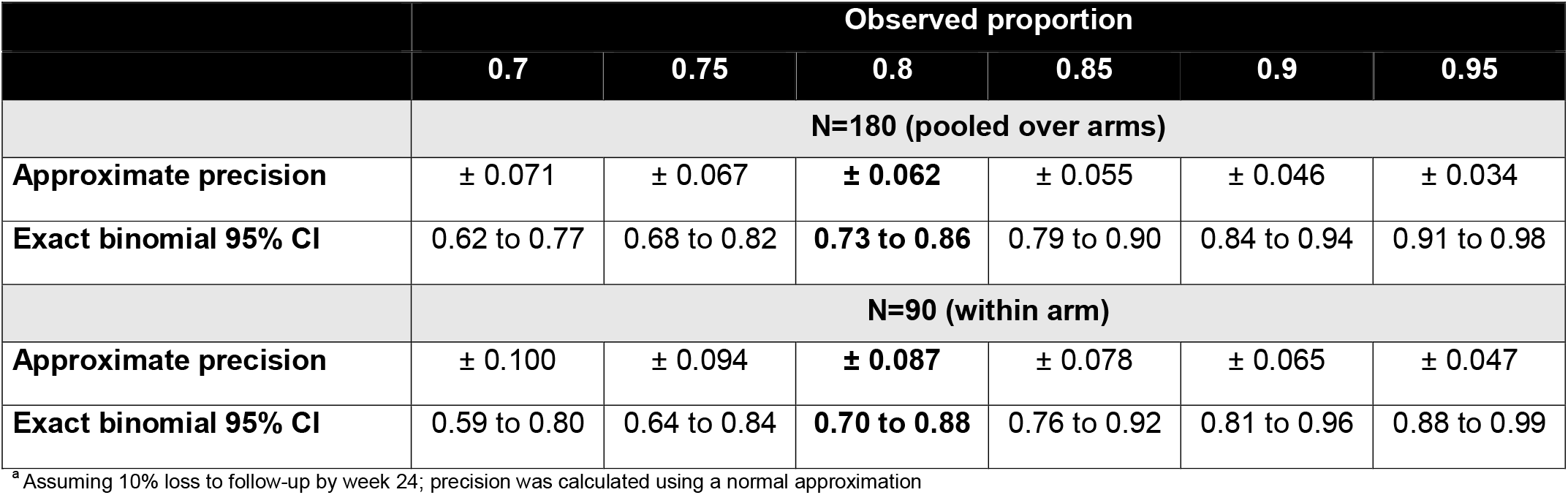
Anticipated Precision of Adherence and Retention Outcomes^a^.

Analogously, we calculated approximate precision and binomial 95% CIs for safety and tolerability with a low proportion of women expected to experience these outcomes. For example, for an observed proportion of 10% experiencing a safety event, we have approximate precision of ±6.5%, separately within each arm. If the observed proportion of women experiencing a tolerability event is 5%, we have approximate precision of ±4.7%, within each arm (Table 2). Regarding statistical power, if the true probability of experiencing a primary safety event is 10% in the placebo arm, then 90 enrolled women per arm will provide us with 90% power to detect a difference in probabilities of 20% or greater (Figure 2).

**Figure 2.**
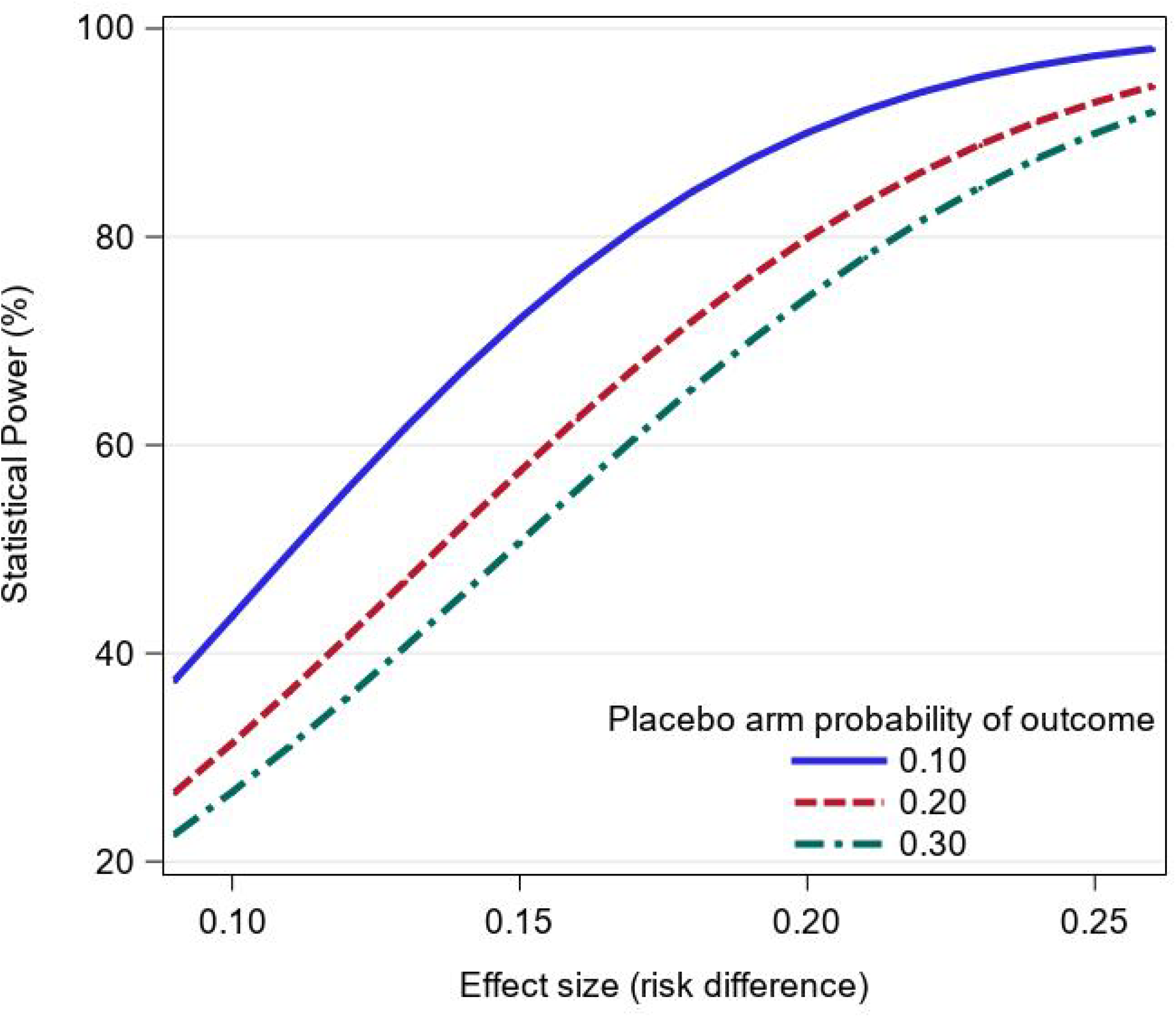
Statistical Power to Detect a Given Difference.

### Ethical Considerations

Participation in the trial is voluntary. Prior to screening, study personnel trained in Good Clinical Practice (GCP) obtain written informed consent from all participants. The study procedures, risks, and benefits are discussed, and women have the opportunity to ask questions prior to providing consent. The informed consent forms are available in English, Sesotho, and Zulu (the three most commonly spoken languages in South Africa) and are written at a fifth grade reading level. Additionally, study staff with participant-facing roles receive protection of human research participants training prior to participating in any study activities and every 2 years thereafter. Key research staff members complete Good Clinical Practice or Good Clinical Laboratory Practice training, as applicable.

It is expected that this trial will expose participants to moderate levels of risk. Participants may experience side effects from the use of intravaginal 5FU. Mild side effects include symptoms such as burning, redness, or vaginal discharge. Serious side effects include pain, bleeding or vaginal ulcers. The proposed treatment regimen (5FU once every 2 weeks) has been previously studied in other settings where women reported mild, grade 1 side effects related to the cream use (e.g., irritation, spotting). However, none of the women enrolled in previously published studies using 5FU once every 2 weeks reported side effects noted to be grade 2 or higher. Additionally, none of the side effects experienced by women in prior studies were reported to interfere with usual activities.

Additional risk associated with study participation include the known risks of clinical procedures such as cervical biopsy and LEEP. Cervical biopsies are minimal risk, but women may experience brief cramping and discomfort, and, occasionally, a small amount of bleeding. Bleeding from the biopsy site is be managed by applying an iron-containing substance (such as Monsel’s solution). There is also an extremely rare risk of fever, infection, and pelvic pain following a diagnostic cervical biopsy.

LEEPs require administration of local anesthetic and may also include cramping and discomfort during the procedure. As bleeding from the LEEP site is controlled with either Monsel’s solution or silver nitrate, a dark-colored discharge is expected after the procedure. LEEPs are also associated with a small risk of fever, infection, pelvic pain, and possible preterm birth in the future. All LEEPs proposed during the study are clinically indicated. Participants will not incur any additional risks beyond standard clinical care.

Collecting lower genital tract samples such as, cytology, cervical swabs, vaginal swabs, and CVL are all minimal risk procedures. Participants may experience mild discomfort from the speculum exam and spotting or light bleeding after collection. The risks associated with blood specimen collection include pain and bruising at the venipuncture site, as well as lightheadedness, dizziness, or, rarely, fainting. There is also an extremely rare risk of infection at the venipuncture site.

Finally, the loss of confidentiality is associated with data collection and storage. At each step in the study, we protect participant privacy and confidentiality to reduce these risks. All study procedures are conducted in private. Data are be stored in secured, locked cabinets and on password-protected computers.

There are no direct benefits of participation in this trial. However, women may benefit from enhanced health education and close clinical monitoring. The knowledge obtained will help to guide future research on and implementation of combination treatment for CIN2/3 in WWH.

The trial is externally monitored by a team of experience Clinical Research Associates (CRAs) at FHI Clinical. It is also reviewed annually by an independent Data Safety and Monitoring Committee (DSMC) convened by the Lineberger Comprehensive Cancer Center at the University of North Carolina at Chapel Hill. The DSMC assesses recruitment, accrual, retention, and safety. The committee also considers new data that may become available, including scientific or therapeutic developments that have an impact on participant safety or the ethics of the trial. The DSMC also has the authority to enjoin enrollment or stop all study activities for reasons of participant safety.

## DISCUSSION

Although the introduction of ART has reduced the incidence of some cancers (e.g., Kaposi’s sarcoma, non-Hodgkin lymphoma) among individuals living with HIV, the role of ART in reducing the incidence of cervical cancer and its precursors remains unclear.^66,67^ As increasing numbers of WWH initiate ART in LMICs such as South Africa, longer life expectancies are anticipated to lead to a rising burden of chronic diseases – including cervical cancer – in this population.^68,69^ WWH in LMICs face additional burdens associated with limited access to cervical cancer prevention measures – vaccination, screening, and surgical treatment of cervical precancer.

In preparation for a large-scale trial to test the effectiveness of combination treatment for CIN2/3 in WWH, we are conducting Phase 2b feasibility trial of LEEP combined with intravaginal 5FU. Given the low cure rates achieved with surgery alone among WWH with CIN2/3,^13,14^ a treatment strategy that combines surgery (excision or ablation) with an adjuvant therapy, such as a self-administered intravaginal medication, would be highly impactful on cervical disease persistence/recurrence and improve long-term cancer risk among WWH.

## Data Availability

All data produced in the present work are contained in the manuscript. This is a protocol paper.

## Notes

**Funding:** CJC and LR are funded by the United States National Institutes of Health (NIH) to study combination treatment approaches for cervical precancer in women living with HIV (R01 CA250850). Trainee support for NST was provided by NIH (T32 HD075731). Additional funding for this work was provided by a UNC Lineberger Comprehensive Cancer Center Developmental Award supported in part by a Cancer Center Core Support Grant from NIH (P30 CA016086). KRM, JRK, and CL also received support from the UNC Center for AIDS Research (P30 AI050410). The NIH was not involved in the design or conduct of the study, or in our decision to submit the trial protocol for publication.

**Conflict of interest:** The authors declare no commercial or other financial conflicts of interest.

### Competing Interest Statement

The authors have declared no competing interest.

### Clinical Trial

NCT05413811

### Funding Statement

CJC and LR are funded by the United States National Institutes of Health (NIH) to study combination treatment approaches for cervical precancer in women living with HIV (R01 CA250850). Trainee support for NST was provided by NIH (T32 HD075731). Additional funding for this work was provided by a UNC Lineberger Comprehensive Cancer Center Developmental Award supported in part by a Cancer Center Core Support Grant from NIH (P30 CA016086). KRM, JRK, and CL also received support from the UNC Center for AIDS Research (P30 AI050410). The NIH was not involved in the design or conduct of the study, or in our decision to submit the trial protocol for publication.

