## Supplemental File 1 for "ACT 2 Clinical Trial Design: Acceptability and Feasibility of Combination Treatment for Cervical Precancer Among South African Women Living with HIV"

### **How to apply** the 5FU/Placebo Cream

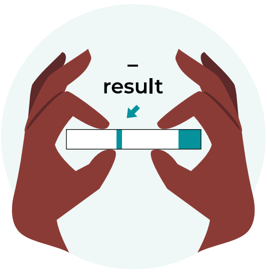


**01**

Take a pregnancy test and **make sure the results are negative before** applying the cream. Record the result in your study diary.

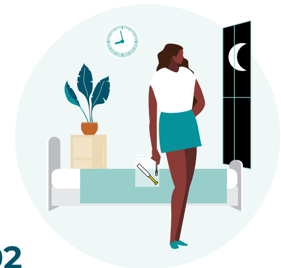

**02**

**Apply the cream at night,** right before you go to bed.

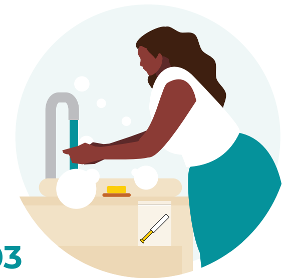

**03**

Wash your hands with soap and water and **remove the pre-filled applicator from its packaging.**

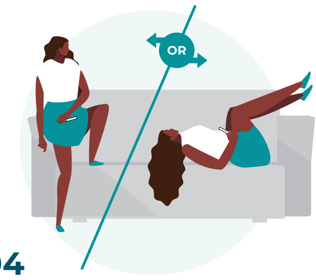


**04**

**Position your body to insert the cream.** Stand with one foot raised or lay on your back with your knees bent and legs shoulder width apart.

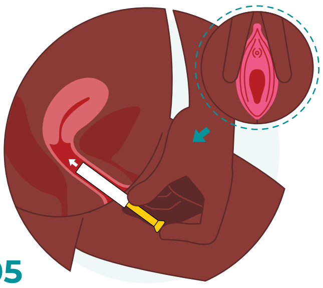

**05**

**With one hand, separate the lips of your vagina. Use the other hand to insert the applicator into your vagina.** Relax your body. This will make it easy to insert the application. It will not be painful.

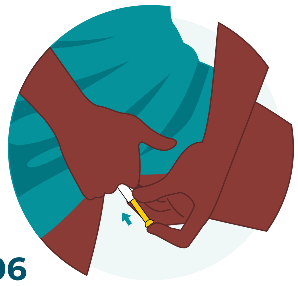

**06**

**When the applicator is in, press down on the plunger-end.** Press until the plunger stops moving and all the cream is inserted.

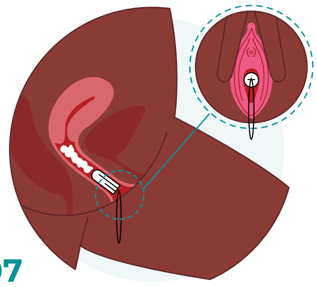

**07**

After inserting the cream, **place a tampon in your vagina to keep the cream from spilling out.**

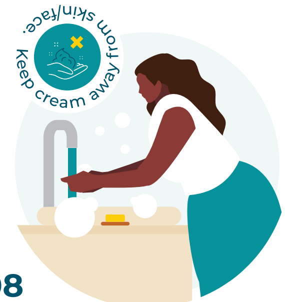


**08**

Wash your hands with soap and water again. This is important to **avoid getting cream on your skin, eyes, mouth, or other body parts.**

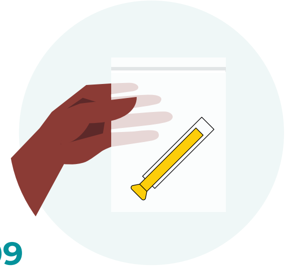

**09**

**Place the used applicator into a plastic bag.** You must return this to the study clinic.

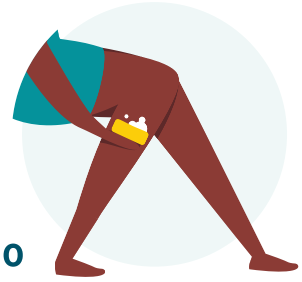

**10**

**In the morning, remove the tampon before you bathe.** Be sure to wash areas of skin that the cream touches.

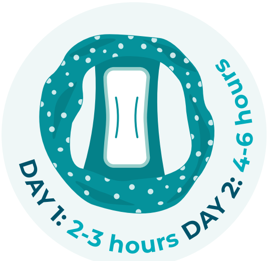


**11**

**Wear a panty liner for 2 days after using the cream. Day 1:** change the panty liner every 2-3 hours. **Day 2:** change the panty liner every 4-6 hours. Continue use after day 2 if it makes you feel comfortable.

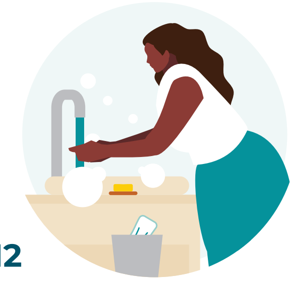

**12**

**Remember to wash your hands with soap and water every time** you change the panty liner.

### **How to use** the 5FU/Placebo Cream

##### What is the cream?

**You insert the cream into your vagina using an applicator.** *The cream should not touch your skin – this will cause irritation.*

##### What is the applicator?

**This is a tube that is filled with the cream.** You use the applicator to insert the cream into your vagina.

##### 5FU or Placebo?

**Some applicators are filled with 5FU cream.** This cream has medicine in it. *We are studying how the medicine works and your feelings about it.*

**Some applicators are filled with placebo cream.** This cream is almost the same as the 5FU cream, except it doesn't have medicine in it. *This is helpful for us to study how well the medicine works between two groups.*

### **Do's** and **Don'ts**

Use **2 forms of birth control during the study.** 5FU can be harmful to pregnancy.

**Do not** use the cream if your pregnancy test is **positive.**

**Do not** have sex *(oral/ vaginal/anal)* for 48 hours after applying the cream.

**Do not** use vaginal washes or herbs.

**Do not** use the cream if you **have your period.** Wait until your period has ended.

**Do not** breastfeed while participating in this study.

**Avoid too much sunlight.** Wear sunblock and a hat, and limit sun exposure to just one hour. Don't use tanning beds.

---

##### **Did you notice** any vaginal irritation?

Apply a small amount of barrier cream, *like Vaseline*, to the outer vaginal area. This will help prevent irritation from the cream. If you feel pain, take paracetamol / ibuprofen.

---

##### **Did you forget** to use the cream?

**Tell the study team.** We will help to adjust your study calendar.

---

##### **Stop using** the cream and notify the study team if you experience:

Abdominal pain, bloody diarrhea, vomiting, fever, chills, mouth sores, vaginal sores, any abnormal bleeding, redness, swelling, numbness, peeling of the skin on the palms and soles, shortness of breath, or hair loss.

**Cervical Cancer Screening and Treatment Clinic**
+27 (0) 11 276 8800 | +27 (0) 72 311 3949 | +27 (0) 72 195 0693
+27 (0) 72 194 9107 | **www.chru.co.za**

5FU Instructional Card v2 27 March 2023
