## Supplemental File 2 for "ACT 2 Clinical Trial Design: Acceptability and Feasibility of Combination Treatment for Cervical Precancer Among South African Women Living with HIV"

#### Statistical Analysis Plan

Katie Mollan, PhD

Principal Biostatistician, University of North Carolina at Chapel Hill

Jessica Keys, PhD

Data Analyst/Manager, University of North Carolina at Chapel Hill

Carla Chibwesha, MD, MSc

Principal Investigator, University of North Carolina at Chapel Hill; Clinical HIV Research Unit at Helen Joseph Hospital, Johannesburg, South Africa

Lisa Rahangdale, MD, MPH

Co-Principal Investigator, University of North Carolina at Chapel Hill

**Version 1.0:** 27 February 2024

**Version 1.1** (minor edits): 25 April 2025

### 1 Introduction

This statistical analysis plan (SAP) details the statistical procedures that address the study objectives specified in **protocol version 4** (21 April 2023) of “ACT 2: Acceptability and Feasibility of Combination Treatment for Cervical Precancer among South African Women Living with HIV”. Analysis plans for any sub-studies not addressed in the protocol, and for additional analyses not anticipated prior to study completion, will be developed as separate documents. Note: Study-specific definitions are provided in **Appendix A**.

#### 1.1 Acronyms

| | |
|---|---|
| ART | Antiretroviral therapy |
| 5FU | 5-fluorouracil |
| CI | Confidence interval |
| CIN | Cervical intraepithelial neoplasia |
| CVL | Cervicovaginal lavage |
| DLT | Dose-limiting toxicity |
| HIV | Human immunodeficiency virus |
| HPV | Human papillomavirus |
| hrHPV | High-risk HPV |
| IQR | Interquartile range, displayed as: $25^{th}$, $75^{th}$ percentile |
| ICC | Intra-class correlation |
| ITT | Intention to treat |
| KM | Kaplan-Meier |
| LEEP | Loop electrosurgical excision procedure |
| RNA | Ribonucleic acid |
| SAP | Statistical analysis plan |
| SD | Standard deviation |
| UVI | Ultraviolet inspection |

### 2 Study Objectives and Summary

**Protocol title:** ACT 2: Acceptability and Feasibility of Combination Treatment for Cervical Precancer among South African Women Living with HIV

**Design:** A double-blind, randomized placebo-controlled feasibility to test the hypothesis that combination treatment for CIN2/3 (i.e., surgical excision followed by adjuvant low-dose topical 5% 5FU) will be safe and well tolerated, with participants adherent to least 6 of 8 doses (75%) of the 5FU cream. HIV-infected women will undergo LEEP and those whose LEEP histology confirms CIN2/3 will be randomly assigned (1:1) to receive 8 doses of intravaginal 5FU or placebo cream once every 2 weeks (5FU/placebo use will begin at week 4).

**Population:** Adult women living with HIV in South Africa who are on ART and are LEEP eligible

**Study groups:** Randomization arm:

- Intervention: 8 doses of intravaginal 5FU
- Control: 8 doses of placebo cream

**Sample size:** We plan to consent and screen approximately 225 women in order enroll and randomize 180 HIV-infected women with CIN 2/3 on LEEP histology.

- Approximately n=90 randomly assigned to active 5FU cream
- Approximately n=90 randomly assigned to placebo cream

**Follow-up:** Participants will be followed for 24 weeks to assess acceptability and feasibility (safety, tolerability, adherence, and retention) of the intervention.

**Study site:** Participants will be enrolled from Themba Lethu HIV Clinic at Helen Joseph Hospital in Johannesburg, South Africa

**Clinical Trial ID:** NCT05413811, clinicaltrials.gov

#### 2.1 Summary of changes to the study protocol(s)

The ACT2 study opened to enrollment in March 2023 under protocol version 3. At protocol version 4 (rolled-out in September 2023), the study team revised the study inclusion criteria as follows:

- Participants are required to be on ART for at least 60 days prior to enrollment. Previously under protocol version 3, the requirement was ART for at least 90 days prior to enrollment.

- LEEP eligibility was expanded to allow for cervical biopsy demonstrating CIN2/3 *or* high-grade (HSIL) cytology on Pap smear within the preceding 12 months. Previously under protocol version 3, cervical biopsy demonstrating CIN2/3 within the preceding 120 days was required for eligibility.

#### 2.2 Study objectives

The following study objectives are from the Protocol (section 1.0):

Our primary objective is:

- To determine the acceptability and feasibility of combination treatment of CIN2/3 (surgical excision followed by adjuvant 5FU) among HIV-infected women.

  <u>Acceptability</u> will be assessed through a participant questionnaire administered at mid-line and end-line.

  <u>Feasibility</u> (safety, tolerability, adherence, and retention) will be assessed over the 24-week study period.

Our secondary objectives are related to the efficacy of combination treatment for CIN2/3.

At study week 24, we will assess:

- Regression of cervical disease to CIN1 or normal histology
- Clearance of the high-risk HPV genotype(s) identified in baseline cervical PreservCyt samples

Our exploratory objectives are:

- Frequency and magnitude of genital HIV-1 shedding and measures of local immune activation. At each timepoint, we will quantify HIV-1 RNA levels and determine expression of innate (IFNα2), immune mediating (IFNγ, IL-10, IL-12), and pro-inflammatory (IL-1α,-1β,-6,-8, MIP-1α, TNFα) cytokines in CVL in each study arm.
- Clearance of high-risk HPV genotype(s) identified in baseline cervical tissue samples.

### 3 Sample Size Considerations

*The following text is from Protocol section 7.2*: We will enroll 180 HIV-infected women in our feasibility trial. We expect grade 2 or higher safety AEs and tolerability events to be rare (Rahangdale 2014), thus our sample size calculations first addressed acceptability and adherence for determining the sample size, while also considering the anticipated precision for the safety and tolerability endpoints.

Acceptability: To estimate the proportions of women reporting an 80% or higher acceptability score, overall and in each arm, we used the normal approximation confidence limit approach and calculated an exact Clopper-Pearson binomial 95% CI as a sensitivity approach. We assume 80% of women will achieve this acceptability threshold at weeks 10 and 24, and thus enrolling 180 women into our study will provide an anticipated margin of error of ±8.7% within arm:

| | **Observed proportion** | | | | | |
|---|---|---|---|---|---|---|
| | **0.7** | **0.75** | **0.8** | **0.85** | **0.9** | **0.95** |
| | **N=180 (pooled over arms)** | | | | | |
| **Approximate precision** | ± 0.071 | ± 0.067 | **± 0.062** | ± 0.055 | ± 0.046 | ± 0.034 |
| **Exact binomial 95% CI** | 0.62 to 0.77 | 0.68 to 0.82 | **0.73 to 0.86** | 0.79 to 0.90 | 0.84 to 0.94 | 0.91 to 0.98 |
| | **N=90 (within arm)** | | | | | |
| **Approximate precision** | ± 0.100 | ± 0.094 | **± 0.087** | ± 0.078 | ± 0.065 | ± 0.047 |
| **Exact binomial 95% CI** | 0.59 to 0.80 | 0.64 to 0.84 | **0.70 to 0.88** | 0.76 to 0.92 | 0.81 to 0.96 | 0.88 to 0.99 |

*[a]Assuming 10% loss to follow-up by week 24; precision was calculated using a normal approximation*

Adherence: To estimate the proportions achieving adherence overall and within each arm, we used the normal approximation confidence limit approach and calculated an exact Clopper-Pearson binomial 95% CI as a sensitivity approach. We assumed that 80% of women would achieve adequate adherence (i.e., 6 of 8 doses). To measure this proportion with a margin of error <10% (anticipated ±8.7%) within arm, we will enroll 180 women into our study.

Retention: we assume that at least 90% of women will be retained in study follow-up, thus our precision and power calculations for other endpoints assumed 10% attrition over the course of the study. If we observe 90% retention over 24 weeks the 95% CI precision will be (0.85, 0.94) pooled over arms and (0.82, 0.95) within arm.

Safety and tolerability: Analogously, we calculated approximate precision and exact binomial 95% CIs for safety and tolerability with a low proportion of women expected to experience these outcomes. For example, for an observed proportion of 10% experiencing a safety event and 90% remaining free of a safety event, we have approximate precision of ±6.5%, separately within each arm. If the observed proportion of women experiencing a tolerability event is 5%, we have approximate precision of ±4.7%, within each arm. Regarding statistical power, if the true probability of experiencing a primary safety event is 10% in the placebo arm, then 90 enrolled women per arm will provide us with 90% power to detect a difference in probabilities of 20% or greater (i.e., probability of a primary safety event of 30% or higher in the 5FU arm versus 10% in the placebo arm).

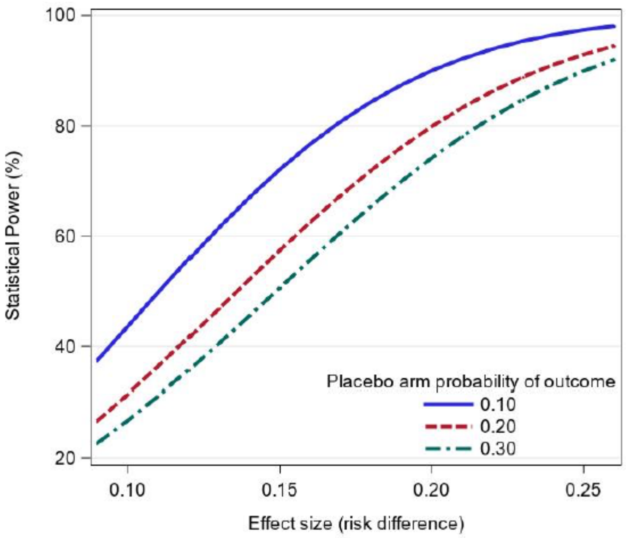

Secondary outcomes: This trial will provide important estimates for future studies. If we assume that regression of cervical disease to CIN1 or normal by week 24 will be achieved by 70% and 90% of women in the placebo and 5FU arms, respectively (Rahangdale 2014), then 90 women enrolled per arm will provide an anticipated margin of error of ±10% for the placebo arm and ±6.5% for the intervention arm. Furthermore, if the true probability of experiencing regression of disease to CIN1 or normal by week 24 is 70% in the placebo arm, then 90 women enrolled per arm provides 90% power to detect of difference in probabilities of 20% or greater (i.e., probability of achieving regression to CIN1/normal of 90% or higher in the 5FU arm versus 70% in the placebo arm).

Background data on clearance of hrHPV genotype(s) are unavailable for this setting. However, for example, assuming that all study participants have at least one hrHPV genotype at baseline, if the percentage of participants who achieve genotype-specific hrHPV clearance by week 24 is 60% in the placebo arm and 80% 5FU arm (Rahangdale 2014), then 90 women enrolled per arm will provide an anticipated margin of error of ±10.7% for the placebo arm and ±8.7% for the intervention arm.

### 4 Randomization and Masking Procedures

We will conduct a double-blind, randomized placebo-controlled trial; the participants, site staff, data collectors, data managers, and study investigators will be masked to the treatment assignment. At study week 4, women with CIN2/3 confirmed by LEEP histology will be randomly assigned 1:1 to 5FU or placebo, and will receive a total of 8 doses of intravaginal 5FU or placebo (one dose every 2 weeks). The 5FU/placebo treatment will begin at week 4 once women have healed from their LEEP. This is a single site study with no stratification factors. The study will enroll and randomize approximately 180 HIV-infected women with CIN 2/3 (*n*=90 per randomization arm).

The randomization schedule (i.e., allocation sequence) was generated by the study statistician in SAS version 9.4 using a random number generator and a blocked randomization design. The block size(s) will be kept concealed from the study team until study conduct is complete. Randomization IDs will be assigned in a concealed manner using a central, web-based REDCap utility. Unmasked study pharmacist will use the randomization schedule to provide the assigned 5FU or placebo treatment. Additional details regarding randomization and masking, including a list of unmasked personnel, can be found in the ACT 2 Randomization Plan Document.

### 5 General Statistical Considerations

Descriptive statistics will be used to (a) delineate the overall study sample that was recruited from a population of adult women living with HIV in South Africa who are on ART and are LEEP eligible, and (b) assess for clinically meaningful imbalances between the randomization arms. Appropriate descriptive statistics will be used throughout, n (%) for categorical data and median (25th, 75th percentile), mean (SD) and min-max for continuous measures. Testing and confidence intervals (CIs) will be two-sided and at a 95% confidence level (i.e., alpha 0.05) with no adjustment for multiplicity, unless stated otherwise.

CONSORT diagram: We will construct a flow diagram of study participants starting at screening. Refer to the Eldridge (2016) reference (pg. 20) for an example of a flow diagram of a parallel randomized pilot trial. We will describe the number of participants screened, enrolled and randomized, and will summarize reasons for any exclusions from each analysis cohort.

Chance imbalance: We anticipate that our measured baseline covariates will be balanced between the arms by randomization. However, if there is considerable chance imbalance, we may conduct a sensitivity analysis applying inverse probability of treatment weights (IPTWs) to construct an analysis cohort with balance in key baseline characteristics that are related to the outcome. A doubly robust approach would also be suitable, if feasible given our sample size (n=90 per arm). The protocol does not address chance imbalance and such an analysis would be a sensitivity approach.

Missing data: Our approach to missing data is provided under each primary and secondary endpoint analysis. Where indicated, if ≥10% of participants in either randomization arm of the analysis cohort are unevaluable we will consider multiple imputation (MI) under a ‘missing at random (MAR)’ assumption to address missing data. However, if the study team decides the missingness is potentially ‘missing not at random (MNAR)’ then alternate strategies will be explored such as nonparametric best and worst case bounds or a tipping point analysis (Liublinska 2014). For analyses where exact CIs are needed to address small sample size considerations, nonparametric best and worst case bounds or tipping point analyses will be applied to address missing data as needed.

To conduct MI, 20 or more imputed datasets will be created using chained equations (MICE; White 2011, Azur 2011) and results will be combined using Rubin’s rule. Missingness patterns will be reviewed prior to finalizing imputation models and each imputation model will be specified before conducting the analysis by randomization arm. Imputation models will include all the covariates in the outcome analysis model and we will consider interaction terms between randomization arm and covariates. Features that are causally related to both the outcome and to missingness will be important for inclusion in the imputation model. As needed, covariate values can also be imputed using MICE, though we anticipate missing baseline data will be uncommon.

Small data: If cell count sizes are small (i.e., for proportions near 0 or 1) then an exact 95% CI approach will be considered as needed for estimating one-sample proportions (Clopper-Pearson CI) and difference in proportions (Chan 1999). Note: exact CI approaches are typically not compatible with applying MI for missing data; in the case of exact inference, we may use nonparametric best and worst case bounds  or tipping point analyses for missing data sensitivity analysis.

Analysis cohorts:

- **ITT**: all participants who are enrolled and randomly assigned to 5FU or placebo (participants who leave the study during the screening phase and without randomization will be excluded from the ITT cohort).

- **Modified ITT**: participants who are eligible, enrolled, randomized, and who receive at least one dose of 5FU or placebo (participants who never start 5FU/placebo will be excluded from the modified ITT cohort).

- **Per-protocol**: Within the modified ITT cohort, those who complete 6, 7, or 8 of the 8 assigned doses of 5FU/placebo and who also attend the week 24 final visit will be considered to have completed study and treatment "per protocol". Those who do not meet this per-protocol definition will have their follow-up right censored at the visit week of their last completed dose of assigned 5FU/placebo.

  As feasible, per-protocol analyses will be conducted where indicated in Primary and Secondary Outcomes section below. If there is a sufficient number of right censoring events, inverse probability of censoring weights (IPCW) will be applied (Hernán 2017) to emulate a per-protocol scenario where all participants who initiate 5FU/placebo go on to complete study follow-up and at least 6 of the 8 assigned treatment doses. If ≥10% of participants in the modified ITT cohort within either randomization arm do not meet the per-protocol definition then we will conduct per-protocol analyses upon request of the PIs. Given the study sample size (n=90 per arm) and scope (feasibility and acceptability), for most the endpoints we anticipate that ITT, modified ITT, or descriptive analyses will be appropriate. The per-protocol definition is based upon the 8-dose 5FU/placebo cream treatment being feasible and acceptable, and seeks to compare 5FU vs. placebo in a setting where all participants who initiate study treatment go on to complete at least 6 of the 8 doses.

### 6 Primary Outcomes

#### 6.1 Acceptability of combination treatment of CIN2/3

Endpoints: (i) Acceptability summary score (range: 0 to 100%)

(ii) Proportion of women who report 80% or higher acceptability summary score

The week 24 analysis will be the primary result with week 10 as a supplemental timepoint.

Cohort: Modified ITT cohort

Exposure: Randomization arm

Measures: Acceptability will be measured at both week 10 and week 24 using a questionnaire with 7 items each using a 5-point Likert scale . Each item will be coded on a range of 0 to 4 for analysis with lowest acceptability = 0, highest acceptability = 4 (**Appendix A**).

Hypothesis: We hypothesize that topical 5% 5FU, a widely available low-cost generic drug, will be acceptable for use as a patient-controlled adjuvant treatment for cervical precancer (CIN2/3) to be self-administered after surgical excision among HIV-infected women.

Analyses:

(i) We will describe the responses to each Likert scale item individually to examine the distribution and missingness of responses at week 10 and 24, separately.

(ii) To compare 5FU vs. placebo, an acceptability summary score will be calculated as a percentage (0 to 100%) of 0 to 28 possible Likert scale points (see details in **Appendix A**), and the distribution of scores will be compared between randomization arms at weeks 10 and 24, separately, using a Wilcoxon rank-sum test accompanied by descriptive statistics. If there is ≥10% missingness in either randomization arm, then MI will be applied to address missing acceptability summary score data.

(iii) In a supplemental descriptive analysis of the observed data, we will calculate item-to-total correlations using Kendall's tau with a corresponding 95% CI to estimate how closely each of the seven Likert scale items (5-level ordinal measure) is correlated to the total acceptability score (continuous measure) at weeks 10 and 24, separately. Measures of agreement may also be considered (e.g., intra-class correlation).

(iv) Lastly, we will estimate the proportion of women in each randomization arm with an acceptability summary score of 80% or higher, with a corresponding Wilson score 95% CI for each randomization arm. We will also estimate the difference in proportions

(5FU vs. Placebo) with a corresponding Wald 95% CI. Missing data will be handled using the approach in analysis (ii) above.

NB: Acceptability of the 5FU cream can only be measured among study treatment initiators (i.e., the modified ITT cohort). For context, we will also describe the number of participants who leave our study prior to 5FU/placebo cream initiation.

#### 6.2 Feasibility (safety, tolerability, adherence, and retention) of combination treatment of CIN2/3

Endpoints:

(i) **Safety**: number and percentage of women experiencing a safety/toxicity event, which for the primary analysis is defined as: any Grade 2 or greater adverse event that that is possibly, probably, or definitely related to study drug, -or- any Grade 1 adverse event of any genital lesion (e.g., blisters, ulcerations, or pustules) that is possibly, probably, or definitely related to study drug. In a supplemental analysis we will assess the number and percentage of women experiencing: any Grade 2 or greater adverse event, -or- any Grade 1 adverse event of any genital lesion (e.g., blisters, ulcerations, or pustules), regardless of relationship to study drug.

(ii) **Tolerability**: the number and percentage of women unable or unwilling to apply at least 4 of 8 doses (50%) of the 5FU/placebo cream due to a dose-limiting toxicity (DLT). A DLT was defined as: (a) any Grade 2 or greater adverse event that that is possibly, probably, or definitely related to study drug and results in an investigator decision to reduce the number of doses or discontinue the study drug, -or- (b) any Grade 1 adverse event of any genital lesion (e.g., blisters, ulcerations, or pustules) that is possibly, probably, or definitely related to study drug and results in an investigator decision to reduce the number of doses or discontinue the study drug.

(iii) **Adherence**: the number and percentage of women who adhere to at least 6 of 8 doses (75%) of 5FU/placebo. Ultraviolet inspection (UVI) will be considered the primary measure, and the applicator counts and self-report will be secondary measures.

(iv) **Retention**: number and percentage of women who are retained in study follow-up over the 24-week study duration (i.e., who attend the week 24 study visit within the allowed study visit window).

Cohorts: (i) Modified ITT
(ii) Per-protocol for safety endpoint only (upon PI request)

Exposure: Randomization arm

Measures: Safety: Adverse event reports including severity grading and relationship to study product will be recorded. Participants will use a dose and symptom diaries to self-report data on cream use and side effects, respectively. During study weeks 4, 6, 10, 18, and 24, participants will undergo pelvic examination and colposcopy. Our trial will adhere to National Institutes of Health (NIH) AE reporting guidelines. We will identify AEs using NCI's Common Terminology Criteria for Adverse Events (CTCAE) version 6.0 and the DAIDS Female Genital Grading Table for Use in Microbicide Studies version 1.0. Safety events can be triggered by graded signs, symptoms, or laboratory abnormalities.

Tolerability: We will collect data on DLTs, 5FU/placebo usage, and study treatment modifications (dose reduction, dose discontinuation).

Adherence: There are three distinct adherence measures: 5FU/placebo applicator counts, ultraviolet inspection (UVI), and self-report questionnaires. Applicators containing 5FU/placebo cream will be placed into a re-sealable bag. Women will be asked to return all used and unused applicators at follow-up visits, and these applicators will be counted and recorded. Applicators will be tested for evidence of vaginal insertion using a UV light. In previous research, the UVI approach had sensitivity of 83% and specificity of 92% and was validated for use in South Africa (Upfold 2017). UVI of used applicators is to be conducted at weeks 4 (for a positive control), 6, 10, 18, and 24. Finally, self-reported data on cream use will collected using a Dose Diary from weeks 4-24.

Retention will be measured using study visit attendance.

Hypothesis: We hypothesize that combination treatment for CIN2/3 (surgical excision followed by adjuvant 5FU) will be safe and well-tolerated compared to a placebo control.

Analyses:

**Safety, tolerability, adherence, and retention** will all be assessed among randomized participants and the time origin for analysis is the date of randomization (i.e., study week 4) unless specified otherwise. For premature study exit prior to the event of interest (i.e., dropout), follow-up will be right-censored at the last respective study measurement. For safety analyses, we will estimate risk within each randomization arm at study week 24 using the Kaplan-Meier (KM) approach with a corresponding 95% CI constructed using Greenwood's variance and a log-log transformation. Follow-up visits will be binned into analysis week windows based on weeks since randomization. The delta method approach will be used with the KM estimator to construct a 95% CI for the risk difference (5FU - placebo) assuming that large sample approximations are tenable. Alternatively, if outcomes are rare (i.e., the number of safety events or non-events is small) then an exact CI for the difference between two independent binomial proportions can be used (Chan 1999); this approach is available in the FREQ Procedure of SAS using the *exact riskdiff* statement with *method=score*.

**(i) Safety analysis**: The at-risk period for safety will begin at study week 4 (randomization and the start of 5FU/placebo cream) and will continue through the last observed study visit where safety was evaluated. We will report the number and percentage of women in each randomization arm experiencing a safety event (see Section 6.2.ii). A KM estimate of safety event probability and a corresponding 95% CI will be constructed for each randomization arm at study week 24, and safety will be compared between the randomization arms using an estimated risk difference (5FU - placebo) with a corresponding 95% CI. Data for the KM analysis will be constructed using an indicator variable for experiencing a safety event ($\delta_s = 1$), and those who do not experience an event will be right censored ($\delta_s = 0$) at their last safety evaluation. The time variable $T$ will be the minimum of: weeks since randomization to first safety event or right censoring (binned into analysis week windows). The same analytic approach will be used to for the supplemental safety endpoint that is regardless of relationship to study drug (see Section 6.2.ii).

**(ii) Tolerability analysis**: The at-risk period for tolerability will begin at randomization and continue through the time of the final dosing visit (i.e., study week 18) or the last tolerability evaluation in the case of premature study exit. If a participant prematurely exits study after experiencing a DLT and before applying at least 50% (i.e. 4 of 8) of the 5FU/placebo cream treatments they will be counted as having a tolerability event at the time of the DLT. We will report the number and percentage of women in each randomization arm unable or unwilling to apply at least 4 of 8 doses of the 5FU/placebo cream due to a DLT. A KM estimate of tolerability event probability and a corresponding 95% CI will be constructed for each randomization arm at study week 18, and tolerability will be compared between the randomization arms using a risk difference (5FU - placebo) with a corresponding 95% CI. Data for the KM analysis will be constructed using an indicator variable for experiencing a tolerability event ($\delta_t = 1$), and those who do not experience an event will be right censored ($\delta_t = 0$) at their last evaluation. The time variable $T$ will be the minimum of: weeks since randomization to first tolerability event or right censoring (binned into analysis week windows).

**(iii) Adherence analysis**: Three sources of adherence data (applicator counts, UVI, and self-report) will be collected and corresponding adherence scores will be estimated from each of the 3 measurement types. Intra-class correlation (ICC) will be used to estimate within-woman agreement between the three data sources (Cassidy 2010).

**UVI will be considered the primary adherence measure and the other two measures will be described as secondary**. For each of the three adherence measures, we will derive a binary endpoint classifying each woman as adherent if she administers at least 6 of 8 doses of 5FU/placebo. Within each randomization arm, we will report the number and percentage of women achieving this adherence threshold with a corresponding Wilson score 95% CI. The percentage of women who achieve adherent dosing will be compared between the randomization arms using an estimated difference in proportions with a corresponding Wald 95% CI. Participants in the analysis cohort who do not achieve 6 adherent doses will be counted as not adherent in our primary analysis (e.g., this may include those who dropout of study follow-up or lose their applicators). Bias induced by missing data is a potential concern for the adherence analyses. If ≥10% of participants in either randomization arm have an unknown UVI adherence outcome due to missing data (e.g., due to dropout or lost applicators) then we will conduct MI for the adherence to each dose and calculate a total number of doses administered (range: 0 to 8) for each imputed dataset. For the imputation model, potential predictors of both missing data and applying the 5FU/placebo cream include, and are not limited to: randomization arm, adherence at non-missing visits, safety events, and DLT.

In a supplemental analysis, we will classify each woman as adherent if she administers 75% or more of her possible 5FU/placebo doses during study follow-up, regardless of early study discontinuation or total number of doses (e.g., the following results would qualify as adherent, ‘doses administered / total doses possible’: 1/1, 2/2, …, 8/8 doses, 3/4, 4/5, 5/6, 6/7, 6/8, 7/8). This analysis will be restricted to observed data while the participant was in study follow-up. If relevant, we may also conduct this supplemental analysis restricted only to those who remained in study follow-up long enough for at least 4 possible doses of 5FU/placebo.

**(iv) Retention analysis**: We will report the number and percentage of women in each randomization arm who are retained in study follow-up at each visit over the 24-week study duration. Week 24 retention will be estimated within each randomization arm with a corresponding binomial Wilson score 95% CI. Week 24 retention will be compared between the 5FU and placebo arms using an estimated difference in proportions and corresponding Wald 95% CI. Among the modified ITT cohort, KM curves by randomization arm will be used to describe weeks from randomization to study dropout or right censoring at the last observed study visit, with dropout event=1 for those who do not complete week 24 and dropout event=0 (right censored) for those who complete study follow-up.

### 7 Secondary Outcomes

#### 7.1 Regression (improvement) of cervical disease to CIN1 or normal

Endpoint: Number and percentage of women who regress to CIN1 or normal by study week 24

Cohorts: (i) ITT
(ii) Modified ITT
(iii) Per-protocol (if feasible and relevant to planning a larger RCT)

Exposure: Randomization arm

Measures: CIN level (normal, 1, 2, 3) will be measured by cervical biopsy at week 24. Consensus review of histology specimens will be performed by pathologists blinded to the randomization arm assignment. Their laboratories are accredited through both the South African National Accreditation System (SANAS) and DAIDS. Proficiency testing is conducted through the Royal College of Pathologists of Australasia Quality Assurance Program. Histology slides will be prepared and stained by a histotechnologist. The pathologist will review all diagnostic and excisional biopsy specimens. Following the 2012 Lower Anogenital Squamous Terminology (LAST) guidelines, objective confirmation of CIN2/3 will be obtained with immunohistochemical staining for p16. Discrepancies will be resolved by consensus review. Similarly, initial screening of the cytology specimens will be performed by a cytotechnologist. All smears will be reviewed by the pathologist. Once again, consultation and consensus review will be used to resolve discrepancies.

Hypothesis: Our central hypothesis is that topical 5% 5FU used as a patient-controlled adjuvant treatment for CIN2/3 to be self-administered after surgical excision will reduce the risk of persistent/recurrent CIN2/3 and progression to cervical cancer among HIV-infected women.

We hypothesize that regression of cervical disease will be more frequent with 5FU treatment compared to placebo.

Analyses: All eligible participants will have CIN2/3 (confirmed by LEEP histology) measured at study week 0 (i.e., baseline for this analysis). We will report the number and percentage of women in each randomization arm who regress (improve) from CIN2/3 at baseline to CIN1 or normal by study week 24 with a corresponding Wilson score 95% CI. Regression of cervical disease will be compared between the randomization arms using an estimated difference in proportions and corresponding Wald 95% CI. We will also describe the week 24 biopsy result using the following groupings: Normal, CIN1, CIN2/3, adenocarcinoma in situ (AIS) or invasive cervical cancer (i.e., adenocarcinoma, squamous cell carcinoma), and Missing. Emphasis will be put on estimation of these secondary outcomes and their feasibility as endpoints in a large RCT; comparisons between the randomization arms will not be considered a definitive results and this endpoint will be clearly stated as secondary in publications and dissemination of results. The main analysis for this endpoint will include those with observed data at week 24 (i.e., we do not plan to impute missing week 24 biopsy data). If there are ≥10% missing data in either randomization arm, then a best and worst case bounds approach can be used to address sensitivity to missing week 24 results.

#### 7.2 Clearance of high-risk HPV genotypes identified at baseline

Endpoint: Number and percentage of women who demonstrate genotype-specific high-risk HPV (hrHPV) clearance between baseline and week 24

Cohorts:
(i) ITT
(ii) Modified ITT
(iii) Per-protocol (if feasible and relevant to planning a larger RCT)

Exposure: Randomization arm

Measures: At baseline (week 0) and week 24, hrHPV genotypes will be measured using HPV DNA testing on cervical PreservCyt samples. All HPV testing will be performed using the GeneXpert system (GeneXpert; Cepheid, Sunnyvale, CA), which employs a cartridge-based real-tome PCR system and reports five separate results: **(a) HPV16, (b) HPV18/45, (c) HPV 31/33/35/52/58, (d) HPV51/59, (e) HPV39/68/56/66.**

Hypothesis: We hypothesize that clearance of all hrHPV genotypes will be more frequent with 5FU treatment compared to placebo

Analyses: Given their diagnosis of CIN2/3, we anticipate that all participants will have at least one hrHPV genotype detected at baseline. We will report the number and percentage of women in each randomization arm who demonstrate genotype-specific hrHPV clearance, defined as having none of the hrHPV types (a)-(e) at week 24 that were originally present at baseline. A corresponding Wilson score 95% CI will be constructed. This composite measure of hrHPV type-specific clearance will be compared between the randomization arms using an estimated difference in proportions and corresponding Wald 95% CI. If the proportion of women who achieve genotype-specific hrHPV clearance is near 0 or 1 in either randomization arm, then an exact score 95% CI for the difference in proportions will be used.

We will also describe the number and percentage of women in each randomization arm with each specific hrHPV type (a)-(e) at baseline and week 24.

The measured hrHPV types will be further described as follows:

(i) Any hrHPV (HPV16/18/45/31/33/35/52/58/51/59/39/68/56/66)
(ii) Any HPV16
(iii) Any HPV18/45 without HPV16
(iv) Any HPV31/33/35/52/58 without HPV16/18/45
(v) Any HPV51/59 or 39/68/56/66 without HPV16/18/45/31/33/35/52/58

We will present data on multiple HPV infections as follows:

(i) 2 or more hrHPV types among (a)-(e)
(ii) 2 or more hrHPV types among HPV16, HPV18/45
(iii) 2 or more hrHPV types among HPV16, HPV18/45, HPV31/33/35/52/58

Emphasis will be put on estimation of these secondary outcomes and their feasibility as endpoints in a large RCT; comparisons between the randomization arms will not be considered definitive results and this endpoint will be clearly stated as secondary in publications and dissemination of results. The main analysis for this endpoint will include those with observed data at week 24 (i.e., we do not plan to impute missing week 24 hrHPV data). If there are ≥10% missing data in either randomization arm, then a best and worst case bounds approach can be used to address sensitivity to missing week 24 results.

### 8 Exploratory Outcomes

Our exploratory outcomes will include frequency and magnitude of genital HIV-1 shedding and measures of local immune activation (i.e., innate, immune mediating, and proinflammatory cytokines) in each randomization arm, over the 24-week study period. We will explore longitudinal changes in the patterns of shedding or cytokine changes. We will also explore clearance of the high-risk HPV genotype(s) identified in baseline cervical tissue samples.

Analysis details and data management plans for exploratory outcomes and measures will be developed prior to conducting exploratory analyses.

### 9 Planned Analyses

#### 9.1 Interim Analyses

We will convene a Data Safety and Monitoring Board (DSMB) to monitor progress of the trial and safety of the participants. The DSMB will review the study protocol and plans for data safety and monitoring. The DSMB will review accrual and safety reports at least once every 6 months and will meet at least annually. DSMB duties will include (a) assessments of recruitment, accrual, retention, and safety; and (b) consideration of new data that may become available, including scientific or therapeutic developments that have an impact on participant safety or the ethics of the trial. For safety, we will assess the number of participants experiencing a DLT. If ≥33% of participants experience a DLT, the DSMB will have authority to enjoin enrollment or stop all study activities for reasons of participant safety. In general, interim analyses of the ACT-2 study will be descriptive. In the case that statistical inference is requested by the DSMB, we will apply a Haybittle-Peto approach to alpha spending at interim analyses (type I error rate $\alpha = 0.001$, and 99.9% confidence intervals). A type I error rate of $\alpha = 0.05$ will be used for the final analysis.

#### 9.2 Final Analysis

The final analysis will be conducted after the last participant has completed the study and will be based on the locked database.

### 10 Changes from the Protocol

With respect to statistical analyses and endpoints, the SAP supersedes the protocol text. Changes in the SAP approach from those stated in the protocol are described below:

**Dose-limiting toxicity (DLT):** In the SAP, we clarified the definition of DLT. See SAP section 6.2.ii for the final definition. Protocol version 4 contained some inconsistencies in the definitions of DLT (protocol sections 6.4 and 8.2).

**Adherence scores**: We removed the composite adherence score and instead will rely on the protocol plan for the case of ICC<0.75 regardless of the observed ICC value. The study investigators decided that assessing UVI, applicator counts, and self-report adherence separately would be better for clinical interpretation as compared to averaging these three measures together. We will still estimate the ICC between measures as a descriptive analysis.

**Missing data:** The SAP was expanded to indicate a missing data approach for each primary and secondary endpoint. While the protocol states "*nonparametric best and worst case bounds will be constructed for missing data sensitivity analyses*" we have updated the SAP to use MI or best-worst bounds accordingly for specific analyses. The tipping point method may also be considered. One advantage of the best and worst bounds approach is it relies on few assumptions and is compatible with exact CI inference; however, these bounds are likely to be wide and potentially difficult for clinical interpretation. A strength of MI is it allows for one summary estimate and CI. However, MI relies on a missing at random assumption and proper specification of the imputation models. Of note, MI is not typically compatible with an exact CI approach and therefore if an exact CI is used then missing data could be handled using best/worst bounds.

### 11 Appendix A: Acceptability summary score

First, each of the following seven Likert scale items (acceptability questions 9-15) will be recoded on a range of 0 to 4, with lowest acceptability = 0 and highest acceptability = 4 for each item. The following items need reverse coded such that “Strongly agree” is equal to 4 to represent the highest acceptability: Q12 (I think the vaginal cream is safe), Q13 (I am confident that I used the vaginal cream correctly), and Q15 (Overall, I had a good experience using the cream).

Then, we will calculate a total score by summing the seven recoded items for 0 to 28 possible acceptability points in total. To ease interpretation, the final acceptability summary score will be calculated as a percentage value (0 to 100%) out of the 28 possible Likert-scale points.

**Adherence and Acceptability Questionnaire** Page **4** of **6**

*For Questions 9-15, please select how strongly you agree or disagree with each of the following statements about the cream you used at home:*

| | |
|---|---|
| **9** | **It was hard to find time to use the cream:**<br>☐ **01-Strongly agree** ☐ **02-Somewhat agree** ☐ **03-Don’t know, no opinion** ☐ **04-Somewhat disagree** ☐ **05-Strongly disagree** |
| **10** | **It was hard to find privacy to use the cream:**<br>☐ **01-Strongly agree** ☐ **02-Somewhat agree** ☐ **03-Don’t know, no opinion** ☐ **04-Somewhat disagree** ☐ **05-Strongly disagree** |
| **11** | **I wanted help from someone to understand how to use the cream:**<br>☐ **01-Strongly agree** ☐ **02-Somewhat agree** ☐ **03-Don’t know, no opinion** ☐ **04-Somewhat disagree** ☐ **05-Strongly disagree** |
| **12** | **I think the vaginal cream is safe:**<br>☐ **01-Strongly agree** ☐ **02-Somewhat agree** ☐ **03-Don’t know, no opinion** ☐ **04-Somewhat disagree** ☐ **05-Strongly disagree** |
| **13** | **I am confident that I used the vaginal cream correctly:**<br>☐ **01-Strongly agree** ☐ **02-Somewhat agree** ☐ **03-Don’t know, no opinion** ☐ **04-Somewhat disagree** ☐ **05-Strongly disagree** |
| **14** | **I was concerned that I would hurt myself while using the vaginal cream:**<br>☐ **01-Strongly agree** ☐ **02-Somewhat agree** ☐ **03-Don’t know, no opinion** ☐ **04-Somewhat disagree** ☐ **05-Strongly disagree** |
| **15** | **Overall, I had a good experience using the cream:**<br>☐ **01-Strongly agree** ☐ **02-Somewhat agree** ☐ **03-Don’t know, no opinion** ☐ **04-Somewhat disagree** ☐ **05-Strongly disagree** |
