## Supplemental File 3 for "ACT 2 Clinical Trial Design: Acceptability and Feasibility of Combination Treatment for Cervical Precancer Among South African Women Living with HIV"

### ACT 2 STUDY PROTOCOL:

**Protocol Version 4, dated 21 Apr 2023**

**Sponsored by:** U.S. National Cancer Institute


**Principal Investigator:** Carla Chibwesha, MD, MSc, FACOG

**Co-Principal Investigators:** Bridgette Goeieman, MBBCh
Lisa Rahangdale, MD, MPH, FACOG

**Co-Investigators:** Malgorzata Beksinska, PhD
Mark Faesen, MBBCh, FCOG
Cecilia Milford, PhD
Masangu Mulongo, MBBCh
Nicholas Teodoro, MD, MPH
Mariza Tunmer, MBChB, FC Rad Onc

#### TABLE OF CONTENTS


| | |
|---|---|
| **PROTOCOL ROSTER** | **3** |
| **PROTOCOL SYNOPSIS** | **4** |
| **1.0 OBJECTIVES** | **6** |
| 1.1 Hypothesis | 6 |
| 1.2 Primary Objectives | 6 |
| 1.3 Secondary Objectives | 6 |
| 1.4 Exploratory Objectives | 6 |
| **2.0 BACKGROUND** | **6** |
| 2.1 Study Disease | 6 |
| 2.2 Topical Treatment Approaches for Cervical Precancer | 7 |
| 2.3 Study Design and Rationale | 7 |
| **3.0 STUDY SETTING** | **8** |
| **4.0 PARTICIPANT SELECTION** | **8** |
| 4.1 Eligibility Criteria | 8 |
| 4.2 Exclusion Criteria | 9 |
| 4.3 Recruitment | 9 |
| 4.4 Proposed Sample Size | 9 |
| **5.0 STUDY INTERVENTION** | **9** |
| **6.0 PROCEDURES** | **10** |
| 6.1 Overview | 10 |
| 6.2 Gynecologic Procedures | 11 |
| 6.3 Laboratory Testing | 11 |
| 6.4 Safety and Tolerability of the Study Intervention | 12 |
| 6.5 Adherence to the Study Intervention | 12 |
| **7.0 STATISTICAL CONSIDERATIONS** | **13** |
| 7.1 Analysis Plan | 13 |
| 7.2 Sample Size and Power | 14 |
| **8.0 ETHICAL CONSIDERATIONS** | **16** |
| 8.1 Human Subjects | 16 |
| 8.2 Risks to Participants | 16 |
| 8.3 Potential Benefits to Participants | 17 |
| 8.4 Financial Compensation | 17 |
| 8.3 Data Safety and Monitoring Board | 17 |
| **9.0 REFERENCES** | **19** |

#### PROTOCOL ROSTER

##### Acceptability and Feasibility of Combination Treatment for Cervical Precancer Among South African Women Living with HIV (ACT 2)


**Principal Investigator:**
**Carla Chibwesha, MD, MSc, FACOG**
Department of Obstetrics and Gynecology
University of North Carolina at Chapel Hill
3009 Old Clinic Building,
NC 27599, USA

Clinical HIV Research Unit
Helen Joseph Hospital, Westdene
Johannesburg 2092, South Africa


**Co-Principal Investigator:**
**Bridgette Goeieman, MBBCh**
Themba Lethu Clinic
Helen Joseph Hospital, Westdene
Johannesburg 2092, South Africa


**Co-Principal Investigator:**
**Lisa Rahangdale, MD, MPH, FACOG**
Department of Obstetrics and Gynecology
University of North Carolina at Chapel Hill
1001 Bondurant Hall
NC 27599, USA


**Biostatistician:**
**Katie Mollan, MS**
Center for AIDS Research
University of North Carolina at Chapel Hill
NC 27599, USA


**Data Manager/Analyst:**
**Jessica Keys, PhD**
Center for AIDS Research
University of North Carolina at Chapel Hill
NC 27599, USA


**Co-Investigators:**
**Malgorzata Beksinska, PhD**
MatCH Research Unit
40 Dr. AB Xuma Street, Suite 1108-9
Durban 4001, South Africa


**Mark Faesen, MBBCh, FCOG**
Clinical HIV Research Unit
Helen Joseph Hospital, Westdene
Johannesburg 2092, South Africa


**Cecilia Milford, PhD**
MatCH Research Unit
40 Dr. AB Xuma Street, Suite 1108-9
Durban 4001, South Africa


**Masangu Mulongo, MBBCh**
Clinical HIV Research Unit
Helen Joseph Hospital, Westdene
Johannesburg 2092, South Africa


**Nicholas Teodoro, MD, MPH**
Clinical HIV Research Unit
Helen Joseph Hospital, Westdene
Johannesburg 2092, South Africa


**Mariza Tunmer, MBChB, FC Rad Onc**
Wits Donald Gordon Medical Centre
21 Eaton Road, Parktown
Johannesburg 2193, South Africa


**PROTOCOL SYNOPSIS**

| | |
|---|---|
| **Title**: | Acceptability and Feasibility of Combination Treatment for Cervical Precancer Among South African Women Living with HIV (ACT 2) |
| **Rationale:** | There are currently no medical therapies recommended to promote the clearance of HPV infection, regression of cervical dysplasia, or treatment of cervical intraepithelial neoplasia (CIN). Development of a noninvasive, patient-controlled treatment modality for CIN is a high priority research area in women's cancer prevention.<br><br>The central hypothesis of the ACT studies is that topical 5-fluorouracil (5FU), a widely available low-cost generic drug, can be used as a patient-controlled adjuvant treatment for cervical precancer (CIN2/3) to be self-administered after surgical excision to reduce the risk of persistent/recurrent CIN2/3 and progression to cervical cancer among HIV-infected women.<br><br>If the proposed combination treatment strategy for CIN2/3 (i.e., surgical excision followed by adjuvant low-dose 5FU) is proven successful, it has the potential to improve long-term outcomes, reducing morbidity and mortality among HIV-infected women who face a substantial lifetime risk of developing cervical cancer. |
| **Overall Objective:** | We will assess the acceptability and feasibility (safety, tolerability, adherence, and retention) of combination treatment for CIN2/3 in clinical trial of intravaginal 5% 5-fluorouracil (5FU)[1] as adjuvant therapy to LEEP. |
| **Study Design:** | We will conduct a double-blind, randomized placebo-controlled **feasibility trial** to test our hypothesis that **combination treatment for CIN2/3** (i.e., surgical excision followed by adjuvant low-dose 5FU) will be safe and well tolerated, with women adherent to least 6 of 8 doses (75%) of the 5FU cream.<br><br>HIV-infected women will undergo LEEP and those whose LEEP histology confirms CIN2/3 will be randomly assigned to receive 8 doses of intravaginal 5FU or placebo cream. Participants will be followed for 24 weeks to assess acceptability and feasibility (safety, tolerability, adherence, and retention) of the intervention. |
| **Outcomes:** | Our primary outcomes are acceptability and feasibility (safety, tolerability, adherence, and retention) of the combination treatment strategy for CIN2/3 in HIV-infected women.<br><br>Secondary outcomes will include the efficacy of the combination treatment strategy. Exploratory outcomes will include frequency and magnitude of genital HIV-1 shedding and measures of local immune activation (i.e., innate, immune mediating, and proinflammatory cytokines) in each study arm. |
| **Setting:** | The ACT studies will be conducted in Johannesburg, South Africa. Participants will be enrolled at the Clinical HIV Research Unit, our study |

| | |
|---|---|
| | site at Helen Joseph Hospital. |
| **Population:** | Our **inclusion criteria** are (a) confirmed HIV-1 infection; (b) age ≥ 18 years; (c) on ART for at least 60 days prior to enrollment; and (d) eligible for LEEP (i.e., those with a cervical biopsy demonstrating CIN2/3 or high-grade (HSIL) cytology on Pap smear within the preceding 12 months).<br>Women will be **excluded** from participation if they are (a) pregnant, breastfeeding, or intend to become pregnant within 180 days of enrollment; (b) have an active sexually transmitted infection (STI; women may participate once treated); (c) have a surgically absent cervix; (d) have a history of anogenital (cervical, vaginal, vulvar, or anal) cancer or a biopsy suspicious for cervical cancer; or (e) have a medical comorbidity that would interfere with study participation. |
| **Procedures:** | We will study the acceptability and feasibility of treating CIN2/3 using LEEP in combination with low dose 5FU.<br>HIV-infected women who are eligible for LEEP will be screened for participation. As part of the trial's screening procedures, women will undergo colposcopy and LEEP at screening (week 0). Women confirmed to have CIN 2/3 on their LEEP histology specimen will then enter the study and be randomly assigned (1:1) to receive 8 doses of intravaginal 5FU or placebo once every 2 weeks (5FU/placebo use will begin at week 4).<br>Women will be asked to return for additional study visits at weeks 6, 10, 18, 24. Study procedures at visit week 6, 10, and 18 include pregnancy testing, pelvic examination, and colposcopy to assess possible adverse events and reinforce adherence. At each of these visits, we will collect cervicovaginal lavage (CVL) samples and for genital HIV viral load and cytokine testing and store a cervical PreservCyt sample and vaginal swab. Additionally, STI testing will be performed at week 10, and if clinically indicated at other timepoints.<br>At week 24 (6 months post-LEEP), women will undergo colposcopically directed cervical biopsies and endocervical curettage (ECC). HPV testing will be performed on cervical PreservCyt samples, and we will collect cervical cytology (Pap smear), CVL, vaginal swabs, and blood samples for CD4+ cell count and plasma HIV viral load. STI testing will also be performed at week 24.<br>Acceptability of the combination treatment approach will be assessed through a questionnaire administered at weeks 10 and 24. Feasibility (safety, tolerability, adherence, and retention) of the approach will be assessed over the 24-week study period. The feasibility analyses will employ an intention-to-treat (ITT) approach. |

#### 1.0 OBJECTIVES

##### 1.1 Hypothesis

The central hypothesis of the ACT studies is that topical 5% 5-fluorouracil (5FU), a widely available low-cost generic drug, can be used as a patient-controlled adjuvant treatment for cervical precancer (CIN2/3) to be self-administered after surgical excision to reduce the risk of persistent/recurrent CIN2/3 and progression to cervical cancer among HIV-infected women.

In the ACT 2 study, a feasibility trial, we hypothesize that combination treatment for CIN2/3 (surgical excision followed by adjuvant 5FU) will be safe and well-tolerated compared to a placebo control, with women adherent to least 6 of 8 (75%) self-applied doses.

As standard surgical treatments for CIN2/3 result in local inflammation and the potential for genital HIV-1 shedding, we further hypothesize that inflammation related to combination treatment will result in transient immune activation and genital HIV shedding, with return to baseline by week 24.

##### 1.2 Primary Objectives

Our primary objective is to determine the acceptability and feasibility of combination treatment of CIN2/3 (surgical excision followed by adjuvant 5FU) among HIV-infected women. Acceptability will be assessed through a participant questionnaire administered at midline and endline. While feasibility (safety, tolerability, adherence, and retention) will be assessed over the 24-week study period.

##### 1.3 Secondary Objectives

Our secondary objectives are related to the efficacy of combination treatment for CIN2/3. At week 24, we will assess (a) regression of cervical disease to CIN1 or normal histology and (b) clearance of the high-risk HPV genotype(s) identified in baseline cervical PreservCyt samples.

##### 1.4 Exploratory Objectives

Exploratory objectives will include the frequency and magnitude of genital HIV-1 shedding and measures of local immune activation. At each timepoint, we will quantify HIV-1 RNA levels and determine expression of innate (IFNα2), immune mediating (IFNγ, IL-10, IL-12), and proinflammatory (IL-1α,-1β,-6,-8, MIP-1α, TNFα) cytokines in CVL in each study arm.

We will also explore clearance of the high-risk HPV genotype(s) identified in baseline cervical tissue samples.

#### 2.0 BACKGROUND

##### 2.1 Study Disease

Cervical cancer remains the second most common cancer among women worldwide, and more than 85% of the global burden of this disease occurs in the developing world. It is well documented that HIV-infected women are at higher risk of HPV infection, with rates as high as 45% - 90%.[2-4] Despite being preventable, cytologic abnormalities, cervical precancer (high-grade cervical intraepithelial neoplasia [CIN2/3]), and invasive cervical cancer also occur more frequently in HIV-infected women.[3,5]

Current management of CIN2/3 is based on surgical therapy alone, which is less effective in HIV-infected women.[6] Surgical therapy for cervical precancer can be excisional (e.g., cold knife conization, loop electrosurgical excision procedure [LEEP]) or ablative (e.g., cryotherapy, thermocoagulation, laser ablation).[7] Excisional treatment requires more surgical skill, but is more appropriate for larger lesions and has the advantage of producing a biopsy specimen for histologic review.[8] Excision is also the standard of care for treatment of CIN2/3 in South Africa.[9]

Although surgical therapy is highly successful in women without HIV,[10,11] cure rates for CIN2/3 are significantly lower in HIV-infected women.[12,13] Three recent African studies (2 conducted by the Clinical HIV Research Unit), enrolling a combined 791 HIV-infected women on antiretroviral therapy (ART), confirm a high risk (19-37%) of recurrent CIN2/3 among women treated with either LEEP or cryotherapy.[14,15] These findings are consistent with a prior U.S. study in which treatment failure occurred in 55% of women with CIN2/3 (N=75) over a 6-year period.[12] By comparison, LEEP treatment failure occurs in only 5-10% of women who are not HIV-infected.[10]

The increase in cervical disease persistence/recurrence among HIV-infected women is thought to derive from residual CIN2/3,[16] re-infection with another hrHPV type, and/or reactivation of underlying hrHPV infection in the genital tract.[17] HIV-infected women could therefore benefit from effective adjuvant therapy following surgery for CIN2/3.

#### 2.2 Topical Treatment Approaches for Cervical Precancer

Multiple observational studies support the efficacy of topical 5FU for treatment of HPV-related diseases (e.g., genital warts, vulvar and vaginal precancer).[18-20] Currently, topical 5FU is also being evaluated in the U.S. for treatment of anal dysplasia (NCT02135419).

Intravaginal 5FU was initially studied in the in U.S. by Maiman et al. as adjuvant therapy for cervical disease in HIV-infected women following surgical treatment for CIN2/3.[21] Women self-applied 2 grams of 5% intravaginal 5FU every 2 weeks for up to 6 months. Histologic outcomes were assessed over 18 months. Participants were relatively immune-compromised; 35% had CD4 counts of <200 cells/mm$^3$ and half were ART-naïve. Compared to women in the observation arm, women treated with 5FU were less likely to experience CIN2/3 recurrence (p=0.014).[21]

More recently, Dr. Rahangdale hypothesized that a similar low-frequency dosing schedule of intravaginal 5FU (2g every 2 weeks) could serve as primary treatment for CIN2 women without HIV.[22] Since the trial enrolled immunocompetent women, therapy was shortened to 16 weeks, consistent with other HPV-related medical management studies.[23-26] Sixty healthy women were randomly assigned to self-apply 5FU every 2 weeks for 16 weeks (8 total doses) versus observation. Compared to observation, women in the 5FU treatment arm were more likely to experience disease regression to CIN1 or normal histology (93% 5FU versus 56% control; p=0.01). Additionally, the composite outcome of histology, Pap smear, and hrHPV result was more likely to be normal in the 5FU treatment group (relative risk: 2.25; 95% CI: 1.05 to 5.09).

With respect to adverse events (AEs), women in the primary treatment trial conducted by Rahangdale et al. only reported grade 1 events (e.g., discharge, burning/irritation, spotting). None of the participants experienced symptoms that caused interference with their usual activities and 83% reported feeling satisfied with the use of 5FU cream.[22] Drs. Chibwesha and Rahangdale have also confirmed the safety of the 8-dose 5FU regimen among HIV-infected women in the U.S.[27]

#### 2.3 Study Design and Rationale

Our overall goal is to reduce the risk of persistent/recurrent CIN2/3 and progression to cervical cancer among HIV-infected women. The ACT studies build on our team's prior research exploring the utility of self-applied intravaginal 5FU as a possible treatment for CIN2/3. Specifically, the ACT studies seek to evaluate the role of a combination treatment approach CIN2/3 in HIV-infected women (i.e., surgical excision followed by adjuvant intravaginal 5FU).

Using a narrative research design, the **ACT 1 study** (which will be completed prior to initiation of this protocol) will assess the **acceptability of the combination treatment approach for CIN2/3**, through focus groups with HIV-infected women and in-depth interviews with male partners and healthcare personnel.

Following this, the **ACT 2 study** (the current protocol) will determine the **acceptability and feasibility of combination treatment of CIN2/3 among HIV-infected women.** In ACT 2, HIV-infected women who are eligible for LEEP will undergo LEEP and those whose LEEP histology confirms CIN2/3 will be randomly assigned to receive 8 doses of intravaginal 5FU cream or placebo cream. Participants will be followed for 24 weeks to assess acceptability and feasibility (safety, tolerability, adherence, and retention) of the combined treatment approach (FIGURE 1).

**Figure 1. Trial Schema**

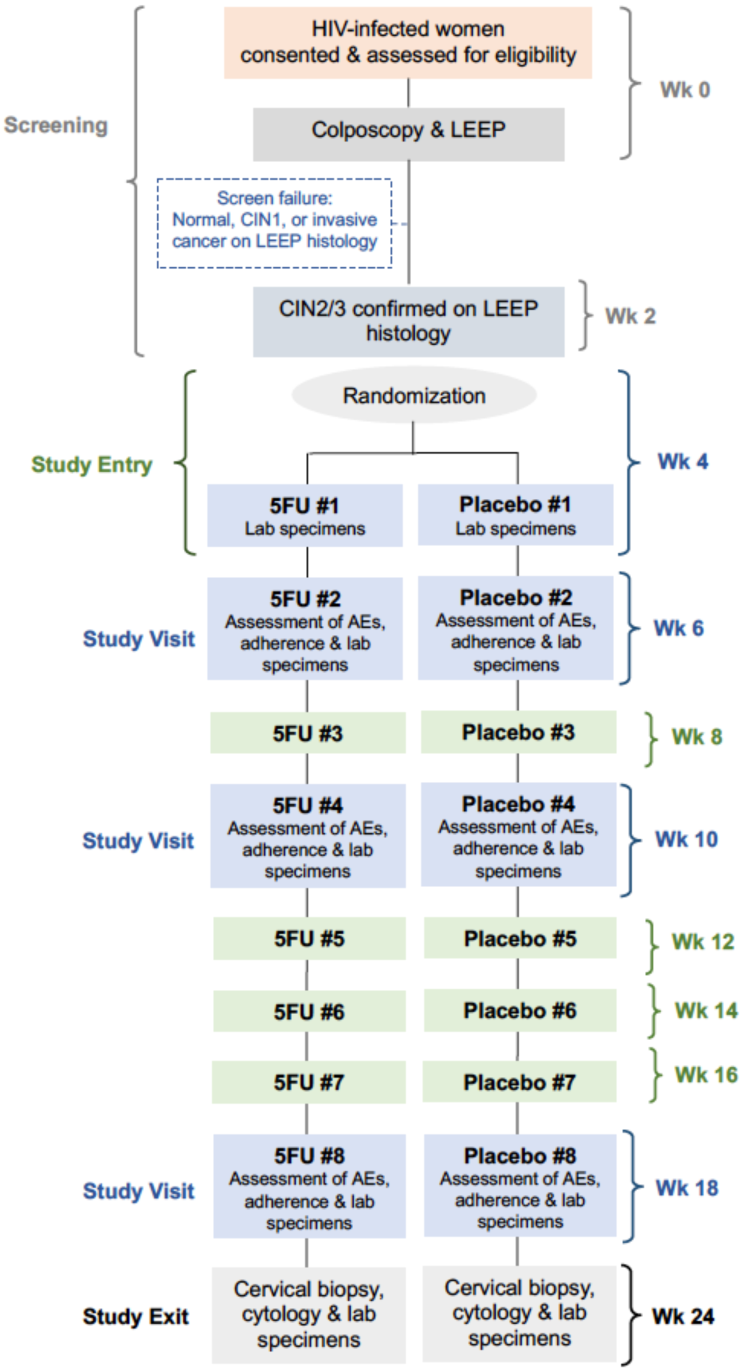


#### 3.0 STUDY SETTING

The ACT studies will be conducted in Johannesburg, South Africa by the Clinical HIV Research Unit (CHRU), a division of Wits Health Consortium. CHRU and its partner NGO, Right to Care, have been national leaders in cervical cancer prevention efforts for over a decade.[14,28-41]

Clinical services and research in this domain currently supported through the **UNC-Wits-Right to Care Partnership for Women's Cancer Prevention**. Co-directed by Drs. Chibwesha and Mulongo, this partnership has provided cervical screening to nearly 100,000 women across South Africa.

Participants for the ACT 2 study will be recruited primarily from **Themba Lethu HIV Clinic (TLC) at Helen Joseph Hospital**.

#### 4.0 PARTICIPANT SELECTION

##### 4.1 Eligibility Criteria

Women who are eligible for LEEP will be invited to learn about the study by a member of our team trained in Good Clinical Practice (GCP). Our inclusion criteria are (a) confirmed HIV-1 infection;

(b) age ≥ 18 years; (c) on ART for at least 60 days prior to enrollment; and (d) eligible for LEEP (i.e., those with a cervical biopsy demonstrating CIN2/3 or HSIL cytology on Pap smear within the preceding 12 months). To be eligible, women should also be willing and able to provide written informed consent.

##### 4.2 Exclusion Criteria

Women will be excluded from participation if they are (a) pregnant, breastfeeding, or intend to become pregnant within 180 days of enrollment; (b) have an active sexually transmitted infection (STI; women may participate once treated); (c) have a surgically absent cervix; (d) have a history of anogenital (cervical, vaginal, vulvar, or anal) cancer or a biopsy suspicious for cervical cancer; or (e) have a medical comorbidity that would interfere with study participation.

Because intravenous 5FU has been shown to be a teratogen in animal models and is classified as a Pregnancy Category X drug, study participants will be required to use dual contraception (condoms plus hormonal contraception, an intrauterine device, implant, or sterilization) during the study.[1] Hormonal contraception is readily available at TLC and 80% of women in a prior TLC studies were using contraception.[14]

##### 4.3 Recruitment

The clinical team will include a brief description of the study as part of their outreach activities and cervical cancer education talks offered in outpatient settings across the hospital, as well as at our referral clinics.

Women who are scheduled to undergo LEEP treatment at Helen Joseph Hospital will be approached about study by a trained member of our team. The study will be explained both verbally and using visual aids in a private location, and women interested in participation will be consented and screened for the study.

##### 4.4 Proposed Sample Size

We plan to consent and screen approximately 225 women in order to enroll and randomize 180 HIV-infected women with CIN 2/3 on LEEP histology. Based on programmatic data from TLC, we anticipate that 15-20% of the 225 women (i.e., approximately 45 women) will have LEEP histology results with CIN1. These women will be considered to have failed screening and will not be enrolled in the study.

We anticipate grade 2 or higher AEs and tolerability issues to be rare.[22] Thus, our sample size of 180 women considers adherence, retention, and acceptability as the main outcomes. To estimate adherence overall and in each arm with adequate precision, we used the normal approximation confidence limit approach and also calculated an exact binomial 95% CI as a sensitivity approach. We assumed that 80% of women will achieve adequate adherence (i.e., at least 6 of 8 doses). To measure this proportion with a margin of error <10% within arm, 180 women will need to be enrolled and randomized. Our calculations also include an estimated 10% loss to follow-up over the course of the study. Additional sample size justifications are provided in Section 7.2.

#### 5.0 STUDY INTERVENTION

We will study the acceptability and feasibility of treating CIN2/3 using **LEEP in combination with adjuvant low dose 5FU.** LEEP will be provided in accordance with the standard of care. 5FU cream (trade name Efudix) will be supplied by the local distributor and held in the CHRU pharmacy. The study placebo cream will be an inert emollient that mimics the consistency and coloring of the 5FU cream. Vaginal applicators pre-filled with 2g of 5FU cream will be distributed to participants to self-apply the cream intravaginally once every 2 weeks for a total of 8 doses. The placebo cream will also be supplied to participants in pre-filled applicators and have identical dosing instructions.

#### 6.0 PROCEDURES

##### 6.1 Overview

HIV-infected women who are eligible for LEEP (i.e., those with a cervical biopsy demonstrating CIN2/3 or HSIL cytology on Pap smear within the preceding 12 months) will be screened for participation. Those wishing to participate will provide written informed consent and be assessed for eligibility, which will include medical history and concurrent medication use, urine pregnancy testing, pelvic examination, STI testing for *C. trachomatis, N. gonnorhea, T. vaginalis* and screening for syphilis.

Eligible participants will undergo colposcopy and **LEEP at their screening visit (week 0)**. The excised cervical tissue will be submitted for histopathology review and immunohistochemical (IHC) staining. Additional baseline specimens collected at the screening visit will include a cervical PreservCyt sample for HPV testing, cervicovaginal lavage (CVL) and for genital HIV-1 viral load and cytokine testing, a vaginal swab for storage, and blood samples for CD4 and HIV-1 plasma viral load (TABLE 1).

**Table 1. Schedule of Events**

| | Week 0 | Week 2 | Week 4 | Week 6 | Week 8 | Week 10 | Week 12, 14, 16 | Week 18 | Week 24 |
|---|---|---|---|---|---|---|---|---|---|
| | Screening A | Screening B | 5FU/P #1 1 month post-LEEP | 5FU/P #2 | 5FU/P #3 | 5FU/P #4 | 5FU/P #5, #6, #7 | 5FU/P #8 | Exit 6 months post-LEEP |
| Demographic questionnaire | X | | | | | | | | |
| STI screen/test | X | | | | | X | | | X |
| Pregnancy test | X | X | X | X | X | X | X | X | X |
| Pelvic exam; colposcopy | X | | X | X | | X | | X | X |
| LEEP | X | | | | | | | | |
| 5FU/Placebo cream | | | X | X | X | X | X | X | |
| Dose diary | | | X | X | X | X | X | X | |
| UVI/Applicator count | | | | X | | X | | X | |
| Symptom diary | | | X | X | X | X | X | X | X |
| Assessment of AEs | | | | X | | X | | X | X |
| Acceptability questionnaire | | | | | | X | | | X |
| Cervical biopsy; ECC | | | | | | | | | X |
| HPV test (cervical sample) | X | | X | | | | | | X |
| Stored cervical sample | | | | X | | X | | X | |
| Cervical cytology | | | | | | | | | X |
| Genital HIV viral load | X | | X | X | | X | | X | X |
| Local immune markers | X | | X | X | | X | | X | X |
| Stored vaginal sample | X | | X | X | | X | | X | X |
| CD4 | X | | | | | | | | X |
| Plasma HIV viral load | X | | | | | | | | X |

Women will return at **week 2** to receive their LEEP histology results and confirmation of eligibility. At **week 4**, those with CIN2/3 confirmed on LEEP histology will be randomly assigned (1:1) to receive 8 doses of intravaginal 5FU or placebo once every 2 weeks. The 5FU/placebo treatment will begin at week 4, once women have healed from their LEEP.

At **week 4**, women will undergo pregnancy testing, pelvic examination, and colposcopy, and will receive step-by-step instructions for 5FU/placebo cream use and a demonstration using a pelvic model. Women will **apply the first dose of cream in the study clinic**. They will also be supplied with pre-filled vaginal applicators of 2g doses of 5FU/placebo cream and asked to apply 2g of the cream at home every 2 weeks (8 doses total). Women will be provided with condoms and urine pregnancy tests to be used prior to applying the 5FU/placebo at home. Additionally, we will collect a cervical PreservCyt sample for HPV testing, CVL for genital HIV-1 viral load and cytokine testing, and a vaginal swab for storage.

Regardless of study arm, women will be asked to return for additional study visits at weeks 6, 10,

18, 24**.** Study procedures planned during the visits at **week 6, 10, and 18** include pregnancy testing, pelvic examination, and colposcopy to **assess possible adverse events**. Visits at week 6, 10, and 18 will assess and reinforce messaging on adherence. At each of these visits, we will collect a cervical PreservCyt sample for storage, CVL for genital HIV-1 viral load and cytokine testing, and a vaginal swab for storage. Additionally, STI screening will be performed at week 10 and if clinically indicated at other timepoints.

At **week 24** (6 months post-LEEP), women will undergo colposcopically directed **cervical biopsies** and endocervical curettage (ECC). At this visit, we will collect a cervical PreservCyt sample for HPV testing, CVL for genital HIV-1 viral load and cytokine testing, a vaginal swab for storage, and blood samples for CD4 and HIV-1 plasma viral load. STI testing will also be performed at week 24.

##### 6.2 Gynecologic Procedures

The research team will perform all gynecologic procedures in their study clinic at Helen Joseph Hospital and will be blinded to study arm assignment.

Pelvic examination and colposcopy: Pelvic examination of the vulva, vagina, cervix, and perineum will include clinical assessment for STIs and AEs. For colposcopy 5 %acetic acid will be applied to the cervix and digital images of the cervical transformation zone will be obtained. These images will be externally reviewed as a quality assurance measure to minimize inter-observer variability in clinical assessment and biopsy location.

LEEP: After paracervical nerve block with injected lidocaine, a wire loop will be used to excise abnormal-appearing tissue in the cervical transformation zone in accordance with the standard of care. Tissue samples will be sent to our contract research laboratory for histologic evaluation and IHC staining.

Cervical biopsies and endocervical curettage: Participants will undergo colposcopically-directed, 4-quadrant biopsies and ECC to adequately evaluate the cervical transformation zone.[42,43] Tissue samples will be sent for histologic evaluation and IHC staining, HPV testing, and HPV genotyping.

Cervical samples: Cervical specimens for HPV testing/storage will be collected using a Cervex-Brush (Rovers Medical Devices, Oss, Netherlands) and stored in PreservCyt solution (Hologic, Marlborough, MA).

Cervical cytology: Liquid-based cytology (LBC) specimens (Pap smears) will be collected in PreservCyt solution (also known as ThinPrep) using a Cervex-Brush.

Cervicovaginal lavage (CVL): A continuous stream of 10mL of phosphate-buffered saline will be aimed directly at and into the cervical opening to bathe the endocervix and ectocervix. The fluid will pool in the posterior fornix and then be aspirated. Samples will be centrifuged, aliquoted, and frozen at -70ºC or below until they are tested.

Vaginal samples: A polyester-tipped swab used to swab the walls of vagina. The swab will be placed into sterile tube spiked with normal saline and then aliquoted and stored at -70ºC or below until tested.

##### 6.3 Laboratory Testing

Cytology and histology: Central review of both cytology and histology specimens will be performed by pathologists blinded to the study arm assignment. Their laboratories are accredited through both the South African National Accreditation System (SANAS).

Histology slides will be prepared and stained by a histotechnologist. The pathologist will review all diagnostic and excisional biopsy specimens. Following the 2012 Lower Anogenital Squamous Terminology (LAST) guidelines,[44,45] objective confirmation of CIN2/3 will be obtained with immunohistochemical staining for p16. Discrepancies will be resolved by consensus review.

Similarly, initial screening of the cytology specimens will be performed by a cytotechnologist. All smears will be reviewed by the pathologist. Once again, consultation and consensus review will be used to resolve discrepancies.

HPV Testing: HPV testing of cervical PreservCyt samples will be performed using the GeneXpert system (GeneXpert; Cepheid, Sunnyvale, CA), which employs a cartridge-based real-time PCR system and reports five separate results: (a) HPV16, (b) HPV18/45, (c) HPV 31/33/35/52/58, (d) HPV51/59, (e) HPV39/68/56/66.

Additionally, as an exploratory analysis, we will examine HPV endpoints from formalin-fixed, paraffin-embedded (FFPE) cervical tissue specimens collected at baseline and endline.[46] This analysis of tissue will be used to assign the causal HPV type in the baseline CIN2/3 lesion and define clearance of the causal type during study follow-up.

HIV-1 RNA viral load testing: CVL specimens stored at -70°C (or below) will be thawed on ice and used to measure HIV-1 RNA on the Abbott RealTime HIV-1 assay (Abbott Molecular, Des Plaines IL).[47-50] Genital HIV shedding will be defined as CVL HIV-1 RNA > 40 copies/mL.

Cytokine testing: CVL specimens stored at -70°C (or below) will be thawed on ice and used to determine expression of innate (IFNα2), immune mediating (IFNγ, IL-10, IL-12), and proinflammatory (IL-1α,-1β,-6,-8, MIP-1α, TNFα) cytokines measured by Luminex technology using Milliplex Human Cytokine Magnetic-bead Kits (Millipore Corporation, Billerica MA).[51,52] To ensure all measurements fall within the standard curve range, 3 randomly selected CVL samples per study arm will be tested in a pilot experiment to determine the optimal dilution. Samples will be tested neat, in duplicate.[49,50]

STI testing: We will use the GeneXpert system (GeneXpert; Cepheid, Sunnyvale, CA) to test for *N. gonnorhea*, *C. trachomatis*, and *T. vaginalis* using vaginal samples collected in accordance with the manufacturer's instructions.[53-56] Rapid testing for syphilis will be performed using Syphilis BD Macro-Vue (Abbott, Abbott Park IL), a screening assay for *T. pallidum* antibodies.[57] Women diagnosed with STIs will receive treatment in accordance with the local standard of care.

Specimen storage: Residual plasma and cervical PreservCyt samples will be aliquoted and stored at -70ºC (or below) for future HPV, DNA methylation, and related biomarker analyses. Vaginal samples will also be aliquoted and stored at -70ºC (or below) for future HPV and microbiome analyses. Lastly, FFPE tissue specimens will be stored at ambient temperature for future HPV, DNA methylation, and related biomarker analyses.

#### 6.4 Safety and Tolerability of the Study Intervention

Our trial will adhere to National Institutes of Health (NIH) AE reporting guidelines. We will identify AEs using NCI's Common Terminology Criteria for Adverse Events version 5.0 and the DAIDS AE Grading Table version 2.1.

Assessment of safety: Participants will use a symptom diary to self-report data on cream use and side effects. During study visits at weeks 4, 6, 10, 18, and 24, participants will undergo pelvic examination and colposcopy. All AEs will be graded using the DAIDS AE Grading Table.

Assessment of tolerability: We will collect data on toxicities as outlined above and report dose-limiting toxicities, defined as any AE possibly, probably, or definitely related to the intervention that leads to treatment discontinuation.

#### 6.5 Adherence to the Study Intervention

We will study adherence to intravaginal cream using a combination of data collection methods to limit the potential for recall and social desirability bias often introduced when data collection occurs at study visits rather than in real time.[58-60] Thus, we will measure adherence using 3 previously-validated methods: applicator counts,[61] ultraviolet inspection (UVI),[62] and self-report.[63]

Applicators containing 5FU/placebo cream will be placed into a resealable bag. Women will be asked to return all used and unused applicators at follow-up visits, and these applicators will be counted and recorded.[61] Applicators will be tested for evidence of vaginal insertion using a UV light. The sensitivity and specificity of the UVI approach (previously validated in South Africa) is 83% and 92%, respectively.[62] Finally, self-reported data on cream use will collected using a symptom diary.

### 7.0 STATISTICAL CONSIDERATIONS

#### 7.1 Analysis Plan

Descriptive statistics will be used to (a) delineate the overall study sample that was recruited from a population of South African adult women living with HIV who are on ART and have a clinical indication for LEEP, and (b) assess for clinically meaningful imbalances between the randomly assigned study arms.

Our **primary outcomes** include **acceptability and feasibility** (safety, tolerability, adherence, and retention). **Acceptability** will be measured at weeks 10 and 24 using a questionnaire with 7 Likert scale items. Each item will be coded for analysis as: lowest acceptability = 0, and higher scores will reflect higher reported acceptability. We will describe the responses to each Likert scale item individually to examine the distribution of responses. To compare acceptability of 5FU vs. placebo, a summary score will be calculated as a percentage (0 to 100%) of 0 to 28 possible Likert scale points, and the distribution of scores will be compared between randomization arms at weeks 10 and 24, separately, using a Wilcoxon rank-sum test accompanied by descriptive statistics. We will also calculate an item-to-total correlation to assess how closely each item is correlated to the total acceptability score. We will then estimate the proportion of women who reported 80% or higher acceptability summary score in each randomization arm, and a corresponding 95% CI will be constructed for each study arm. Lastly, we will estimate the difference in proportions (5FU vs. Placebo) with a corresponding 95% CI.

**Safety, tolerability, adherence, and retention** will all be assessed over the 24-week study period. Analyses will employ a modified intention-to-treat (ITT) approach (participants who never start 5FU/placebo will be excluded), and we will also conduct a per-protocol analysis for safety and tolerability. All testing and confidence intervals (CIs) will be two-sided and at a 95% confidence level (i.e., alpha 0.05).

For premature study exit prior to the event of interest (i.e., attrition), follow-up will be right-censored at the last respective study measurement and outcome risk will be estimated using the Kaplan-Meier (KM) approach. The delta method or a bootstrap approach can be used with the KM estimator to construct a 95% CI for the difference in proportions (i.e., risk difference) assuming that large sample approximations are tenable. However, an exact CI for the difference between two independent binomial proportions will be considered if outcomes are rare (i.e., if the number of events or non-events is small).[64] Primary analyses will use a complete-case (missing data excluded) approach, and nonparametric best and worst case bounds will be constructed for missing data sensitivity analyses.[65]

<u>Safety endpoint</u>: We will report the number and percentage of women in each study arm experiencing a grade 2 or higher toxicity, or any grade 1 genital toxicity (blisters, ulcerations, or pustules), that is possibly, probably, or definitely related to the study intervention. A corresponding 95% CI will be constructed for each study arm. Safety endpoints will be compared between the 5FU and placebo arms using an estimated difference in proportions with a corresponding 95% CI. The at-risk period for safety events will begin on study week 4 (at the start of 5FU/placebo cream) and will continue through study week 24 or the last attended safety evaluation in the case of premature study exit.

Tolerability endpoint: We will report the number and percentage of women in each study arm unable or unwilling to apply at least 4 of 8 doses (50%) of the 5FU/placebo cream due to a dose-limiting toxicity (i.e., any AE regardless of grade that is possibly, probably, or definitely related to the intervention that leads to treatment discontinuation); a corresponding 95% CI will be constructed for each study arm. Tolerability endpoints will be compared between the study arms using an estimated difference in proportions with a corresponding 95% CI. The at-risk period for tolerability events will begin on study week 4 and continue through study week 18 (a 14-week duration) or the last tolerability evaluation in the case of premature study exit. If a participant prematurely exits study after experiencing a dose-limiting toxicity and before applying at least 50% of the 5FU/placebo cream treatments she will be counted as a tolerability event.

Adherence endpoint: 3 sources of adherence data (applicator counts, UVI, and self-report) will be collected and the corresponding adherence rates will be reported. Intra-class correlation (ICC) will be used to estimate within-woman agreement between the three data sources.[66] Assuming high agreement (correlation of 75% or higher), a primary composite adherence rate will be computed by averaging the data from all 3 sources (each carrying equal weight). If agreement (ICC) between the data sources is below 75%, UVI will be considered the primary adherence measure and the other 3 measures will be described as secondary.

We will classify a woman as adherent if she administers at least 6 of 8 doses (75%) of 5FU/placebo. Given that 6 of 8 doses will be required to achieve adherence, this endpoint will require both retention to study follow-up and adherence to dosing. We will report the number and percentage of women achieving adherence. A corresponding 95% CI will be constructed for each study arm. Also, this endpoint will be compared between the study arms using an estimated difference in proportions with a corresponding 95% CI.

Retention endpoint: We will report the number and percentage of women in each study arm who are retained in study follow-up over the 24-week study duration (i.e., who attend the week 24 study visit). A corresponding binomial 95% CI will be constructed for each study arm. 24-week retention will be compared between the 5FU and placebo arms using an estimated difference in proportions and corresponding 95% CI. Kaplan-Meier curves by study arm will be used to describe weeks from randomization to the last attended study visit with follow-up censored at the date of last study contact.

**Our secondary outcomes** are related to the **efficacy of combination treatment** for CIN2/3. At week 24, we will assess (a) regression of cervical disease to CIN1 or normal histology and (b) clearance of the hrHPV genotype(s) identified at baseline.

Regression of cervical disease between baseline and week 24: All participants will have CIN2/3 (confirmed by LEEP histology) at baseline, and we will report the number and percentage of women in each study arm who regress to CIN1 or normal by week 24 with corresponding binomial 95% CIs. Regression of cervical disease will be compared between the study arms using an estimated difference in proportions and corresponding 95% CI.

Clearance of hrHPV genotype(s) between baseline and week 24: Given their diagnosis of CIN2/3, we anticipate that all participants will have at least one hrHPV genotype detected at baseline. As a **secondary outcome**, we will consider individual hrHPV genotypes and report the number and percentage of women in each study arm who demonstrate **genotype-specific hrHPV clearance** between baseline and week 24 with corresponding binomial 95% CIs. HPV clearance will be compared between the study arms using an estimated difference in proportions and corresponding 95% CI.

Our **exploratory outcomes** will include **frequency** and **magnitude of genital HIV-1 shedding** and **measures of local immune activation** (i.e., innate, immune mediating, and proinflammatory cytokines) in each study arm, over the 24-week study period. We will also explore longitudinal changes in the patterns of shedding or cytokine changes.

**Figure 2. Statistical Power to Detect a Given Risk Difference**

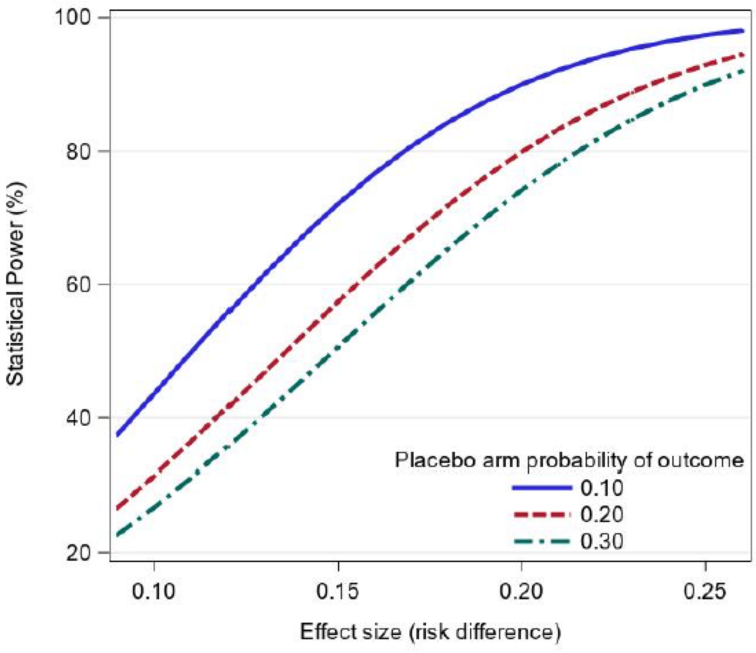


Safety and tolerability: Analogously, we calculated approximate precision and exact binomial 95% CIs for safety and tolerability with a low proportion of women expected to experience these outcomes. For example, for an observed proportion of 10% experiencing a safety event and 90% remaining free of a safety event, we have approximate precision of ±6.5%, separately within each arm. If the observed proportion of women experiencing a tolerability event is 5%, we have approximate precision of ±4.7%, within each arm (TABLE 2). Regarding statistical power, if the true probability of experiencing a primary safety event is 10% in the placebo arm, then 90 enrolled women per arm will provide us with 90% power to detect a difference in probabilities of 20% or greater (i.e., probability of a primary safety event of 30% or higher in the 5FU arm versus 10% in the placebo arm; FIGURE 2).

Secondary outcomes: This trial will provide important estimates for future studies. If we assume that regression of cervical disease to CIN1 or normal by week 24 will be achieved by 70% and 90% of women in the placebo and 5FU arms, respectively,[22] then 90 women enrolled per arm will provide an anticipated margin of error of ±10% for the placebo arm and ±6.5% for the intervention arm. Furthermore, if the true probability of experiencing regression of disease to CIN1 or normal by week 24 is 70% in the placebo arm, then 90 women enrolled per arm provides 90% power to detect of difference in probabilities of 20% or greater (i.e., probability of achieving regression to CIN1/normal of 90% or higher in the 5FU arm versus 70% in the placebo arm).

Background data on clearance of hrHPV genotype(s) are unavailable for this setting. However, for example, assuming that all study participants have at least one hrHPV genotype at baseline, if the percentage of participants who achieve genotype-specific hrHPV clearance by week 24 is 60% in the placebo arm and 80% 5FU arm,[22] then 90 women enrolled per arm will provide an anticipated margin of error of ±10.7% for the placebo arm and ±8.7% for the intervention arm.

#### 8.0 ETHICAL CONSIDERATIONS

##### 8.1 Human Subjects

This protocol will be submitted to the University of Witwatersrand Human Research Ethics Committee (Medical) and the University of North Carolina Institutional Review Board.

Informed Consent: Discussions with prospective participants and informed consent procedures will be conducted in private to protect confidentiality. GCP-trained study personnel will obtain written informed consent from all participants. The study procedures, risks, and benefits will be discussed, and participants will have the opportunity to ask questions prior to providing consent. All materials will be designed at a fifth-grade comprehension level and will be available in English, Sesotho, and Zulu. All versions of the consent form and participant education materials will be approved by the relevant ethics committees prior to study initiation. For illiterate participants, a literate impartial witness will be present during the entire consent process to ensure that all of the relevant information has been provided and the participant voluntarily gives consent.

##### 8.2 Risks to Participants

5FU cream: Participants may experience side effects from the use of intravaginal 5FU. Mild side effects include symptoms such as burning, redness, or vaginal discharge. Serious side effects include pain, bleeding or vaginal ulcers. The proposed treatment regimen (5FU once every 2 weeks) has been previously studied in other settings where women reported mild, grade 1 side effects related to the cream use (e.g., irritation, spotting). However, none of the women enrolled in previously published studies using 5FU once every 2 weeks reported side effects noted to be grade 2 or higher. Additionally, none of the side effects experienced by women in prior studies were reported to interfere with usual activities.

Because systemic 5FU is listed as Food and Drug Administration (FDA) Category X, participants should not become pregnant or breastfeed while using the 5FU cream because the drug could be teratogenic. To minimize this risk, women will be required to use dual contraception (condoms plus hormonal contraception, an intrauterine device, or sterilization) while receiving the study drug.

Invasive cervical cancer: It is extremely unlikely that CIN2/3 will progress to cervical cancer during 24 weeks of study follow-up. However, our study will include close observation with surveillance for cervical disease (i.e., colposcopy) performed at each study visit. Additionally, at the exit visit, all women will undergo cervical biopsy and any cases of CIN2/3 or cervical cancer detected will be referred for immediate surgical management. It should also be noted that any possible disease progression would *not* be related to the study intervention.

Cervical biopsy and LEEP: The risks associated with diagnostic cervical biopsies include brief cramping and discomfort from the punch biopsy and, occasionally, a small amount of bleeding. Any bleeding from the biopsy site will be managed by applying an iron-containing substance (such as Monsel's solution) or a silver-containing compound (such as silver nitrate) for hemostasis and may cause a dark-colored discharge. There is also an extremely rare risk of fever, infection, and pelvic pain following a diagnostic biopsy.

LEEPs require administration of local anesthetic and may also include cramping and discomfort during the procedure. As bleeding from the LEEP site is controlled with either Monsel's solution or silver nitrate, a dark-colored discharge is expected after the procedure. LEEPs are also associated with a small risk of fever, infection, pelvic pain, and possible preterm birth in the future. Once again, it should be noted that all LEEPs proposed during the study are clinically indicated. Participants will not incur any additional risks beyond standard clinical care.

Swab, Pap smear: Swabs and Pap smears are minimal risk procedures. Participants may feel discomfort from the speculum exam and may experience minimal bleeding or spotting after collection of the specimen.

Cervicovaginal lavage: CVL is a minimal risk procedure. Participants may feel discomfort from the saline solution.

Urine collection: There are no known risks of urine collection.

Phlebotomy: Risks associated with blood specimen collection include pain and bruising at the venipuncture site, as well as lightheadedness, dizziness, or, rarely, fainting. There is also an extremely rare risk of infection at the venipuncture site.

Confidentiality: The main risk associated with data collection and storage is loss of confidentiality. At each step in the study, we will protect participant privacy and confidentiality to reduce these risks. All study procedures will be conducted in private. Data will be stored in secured, locked cabinets and on password-protected computers.

##### 8.3 Potential Benefits to Participants

There are no direct benefits of participation in the study. However, the knowledge obtained will help to guide future research on and implementation of combination treatment for CIN2/3 in women living with HIV.

##### 8.4 Financial Compensation

Participants will be reimbursed for their time/travel expenses a rate of R400 per study visit.

##### 8.2 Data Safety and Monitoring Board

We will convene a Data Safety and Monitoring Board (DSMB) to monitor progress of the trial and safety of the participants. The DSMB will include approximately 5 members with experience in statistics, cancer epidemiology, gynecology, and public health that are not otherwise involved in the study. The DSMB will meet prior to study start-up to review the study protocol and plans for data safety and monitoring. Thereafter, the DSMB will review accrual and safety reports at least once every 6 months and will meet annually. DSMB duties will include (a) assessments of recruitment, accrual, retention, and safety; and (b) consideration of new data that may become available, including scientific or therapeutic developments that have an impact on participant safety or the ethics of the trial. For safety, we will assess the number of participants experiencing a dose limiting toxicity (DLT). DLT is defined as Grade 2 or greater toxicity (or Grade 1 toxicity of any genital lesion (blisters, ulcerations, or pustules) that is possibly, probably, or definitely related and lasts for more than 5 days. If $\geq$ 33% of participants experience a DLT, the DSMB will have authority to enjoin enrollment or stop all study activities for reasons of participant safety.
